# From a genomic risk model to clinical trial implementation in a learning health system: the ProGRESS Study

**DOI:** 10.1101/2024.11.03.24316516

**Authors:** Jason L Vassy, Anna M Dornisch, Roshan Karunamuni, Michael Gatzen, Christopher J Kachulis, Niall J Lennon, Charles A Brunette, Morgan E Danowski, Richard L Hauger, Isla P Garraway, Adam S Kibel, Kyung Min Lee, Julie A Lynch, Kara N Maxwell, Brent S Rose, Craig C Teerlink, George J Xu, Sean E Hofherr, Katherine A Lafferty, Katie Larkin, Edyta Malolepsza, Candace J Patterson, Diana M Toledo, Jenny L Donovan, Freddie Hamdy, Richard M Martin, David E Neal, Emma L Turner, Ole A Andreassen, Anders M Dale, Ian G Mills, Jyotsna Batra, Judith Clements, Olivier Cussenot, Cezary Cybulski, Rosalind A Eeles, Jay H Fowke, Eli Marie Grindedal, Robert J Hamilton, Jasmine Lim, Yong-Jie Lu, Robert J MacInnis, Christiane Maier, Lorelei A Mucci, Luc Multigner, Susan L Neuhausen, Sune F Nielsen, Marie-Élise Parent, Jong Y Park, Gyorgy Petrovics, Anna Plym, Azad Razack, Barry S Rosenstein, Johanna Schleutker, Karina Dalsgaard Sørensen, Ruth C Travis, Ana Vega, Catharine M L West, Fredrik Wiklund, Wei Zheng, Profile Steering Committee, IMPACT Study Steering Committee and Collaborators, PRACTICAL Consortium, Million Veteran Program, Tyler M Seibert

## Abstract

**Background:** As healthcare moves from a one-size-fits-all approach towards precision care, individual risk prediction is an important step in disease prevention and early detection. Biobank-linked healthcare systems can generate knowledge about genomic risk and test the impact of implementing that knowledge in care. Risk-stratified prostate cancer screening is one clinical application that might benefit from such an approach.

**Methods:** We developed a clinical translation pipeline for genomics-informed prostate cancer screening in a national healthcare system. We used data from 585,418 male participants of the Veterans Affairs (VA) Million Veteran Program (MVP), among whom 101,920 self-identify as Black/African-American, to develop and validate the Prostate CAncer integrated Risk Evaluation (P-CARE) model, a prostate cancer risk prediction model based on a polygenic score, family history, and genetic principal components. The model was externally validated in data from 18,457 PRACTICAL Consortium participants. A novel blended genome-exome (BGE) platform was used to develop a clinical laboratory assay for both the P-CARE model and rare variants in prostate cancer-associated genes, including additional validation in 74,331 samples from the All of Us Research Program.

**Results:** In overall and ancestry-stratified analyses, the polygenic score of 601 variants was associated with any, metastatic, and fatal prostate cancer in MVP and PRACTICAL. Values of the P-CARE model at ≥80th percentile in the multiancestry cohort overall were associated with hazard ratios (HR) of 2.75 (95% CI 2.66-2.84), 2.78 (95% CI 2.54-2.99), and 2.59 (95% CI 2.22-2.97) for any, metastatic, and fatal prostate cancer in MVP, respectively, compared to the median. When high– and low-risk groups were defined as P-CARE HR>1.5 and HR<0.75 for metastatic prostate cancer, the 220,062 (37.6%) high-risk vs.146,826 (25.1%) low-risk participants in MVP had a 47.9% vs. 14.1%, 9.3% vs. 2.0%, and 3.6% vs. 0.8% cumulative cause-specific incidence of any, metastatic, and fatal prostate cancer by age 90, respectively. The clinical assay and reports are now being implemented in a clinical trial of precision prostate cancer screening in the VA healthcare system (Clinicaltrials.gov NCT05926102).

**Conclusions:** A model consisting of a polygenic score, family history, and genetic principal components describes a clinically important gradient of prostate cancer risk in a diverse patient population and demonstrates the potential of learning health systems to implement and evaluate precision health care approaches.

## INTRODUCTION

Preventive healthcare is moving from a one-size-fits-all approach to more personalized, risk-adapted strategies. Individual risk prediction is an important step for developing tailored strategies for disease prevention and early detection. Risk prediction models can now incorporate larger and more complex arrays of clinical, genetic, environmental, and other risk factors from more diverse populations.^1^ Genomics specifically is increasingly demonstrating its potential to inform risk stratification for several diseases.^2–5^ However, much of this potential clinical utility for disease prevention remains theoretical, absent prospective intervention studies demonstrating improved patient outcomes.

Healthcare systems linked to genomic biobanks thus have the opportunity both to generate knowledge about the clinical validity of genomic risk prediction and to demonstrate the clinical utility of implementing that knowledge in care.^6^ These *genomics-enabled learning healthcare systems* can leverage knowledge-generating infrastructure to determine whether genomics and other novel risk predictors improve the effectiveness of disease screening and prevention within the healthcare system. The result is not only improved care for patients within that system but also potentially generalizable knowledge for patients in other settings.

Prostate cancer screening is one clinical context where a genomics-enabled learning healthcare system approach might be particularly beneficial. Prostate cancer is one of the most heritable cancers, and recent genomic discoveries have characterized the rare and common genetic variation underlying much of this heritability.^7,8^ At the same time, clinical guidelines differ on which patient populations are most likely to experience net benefit from prostate cancer screening, including Black men or those with a family history of the disease.^9–11^ Universal screening with prostate-specific antigen (PSA) reduces prostate cancer mortality but can also overdiagnose indolent disease and lead to unnecessary procedures and treatments.^12–15^ The result is significant variation in prostate cancer screening practices.^16,17^ Clinical consensus is even less developed on whether genotype should play a role in prostate cancer risk stratification, despite the discovery of robust associations between risk and both single rare variants and polygenic scores.^8^

Given this context, genomics-enabled learning health systems can lead the development of genomics-tailored approaches to prostate cancer screening and then evaluate the effectiveness of those approaches in clinical care. This evidence generation is an important step towards the development of clinical guidelines. Here, we describe the development, validation, and clinical implementation of a novel genomics-informed prostate cancer risk model, developed to enable a randomized clinical trial of precision prostate cancer screening in a large national healthcare system (Clinicaltrials.gov NCT05926102).

## METHODS

### Study overview

Figure 1 illustrates our discovery-to-implementation approach. As described in detail below, we used data from a large biobank linked to a national healthcare system to update a prior prostate cancer polygenic score.^18^ We then developed and cross-validated a prostate cancer prediction model based on the combination of that score and family prostate cancer history, now termed the Prostate CAncer integrated Risk Evaluation (P-CARE) model. We further validated the P-CARE model in four external prostate cancer cohort datasets prior to the development and validation of a clinical blended genome-exome (BGE) assay both for the P-CARE model and also for rare prostate cancer-associated monogenic variants. This assay is now being implemented in a randomized clinical trial of genomics-informed prostate cancer screening in a new cohort of patients from the national healthcare system in which the P-CARE model was first developed [the Prostate Cancer, Genetic Risk, and Equitable Screening Study (ProGRESS), ClinicalTrials.gov ID NCT05926102]. The VA Central IRB approved this study (IRBNet 1735869 and 1735136).

**Figure 1:**
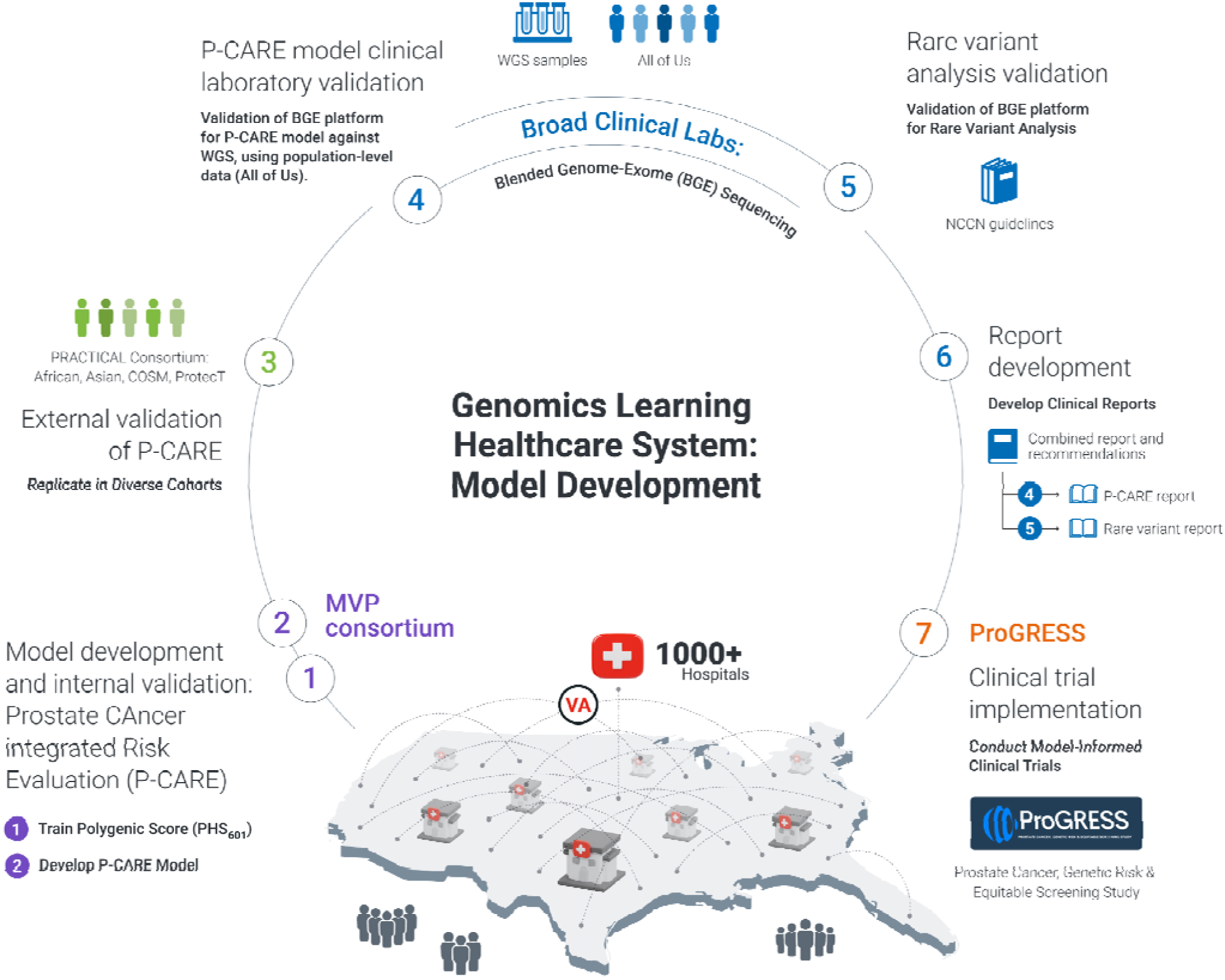
Translating prostate cancer genomic risk discovery to clinical trial implementation. 1) A prostate cancer polygenic score (PHS_601_) is trained in the Million Veteran Program (MVP) biobank of the Veterans Health Administration (VA) using known prostate cancer and other prostate trait-associated loci. 2) The Prostate CAncer Risk and Evaluation (P-CARE) model is developed in MVP from PHS_601_, genetic components, and prostate cancer family history. 3) Both PHS_601_ and P-CARE are replicated in external multiancestry datasets from the Prostate Cancer Association Group to Investigate Cancer Associated rincipal Alterations in the Genome (PRACTICAL) consortium. 4) A blended genome-exome (BGE) platform is validated for the P-CARE model, including imputation, analytic validation of PHS_601_ against whole genome sequencing (WGS) and clinical laboratory validation of P-CARE in All of Us Research Program data. 5) BGE platform is validated for gene panel annotation, filtering, and analysis of rare variants in guideline-informed prostate cancer-associated genes. 6) Clinical P-CARE and rare variant reports with summary recommendations are developed. 7) Clinical laboratory analysis and reporting pipeline is implemented in a pragmatic clinical trial of precision prostate cancer screening across the VA.

### Participants and phenotype definitions

Genotype and phenotype data were analyzed from the following cohorts, described previously.^19–21^ and summarized in **Supplemental Tables 1 and 2**.

### Million Veteran Program

Data from the Million Veteran Program (MVP) were used to update a previous prediction model^18^ to develop the new P-CARE model. MVP is a mega-biobank linked to the national Veterans Health Administration healthcare system of the US Department of Veterans Affairs (VA).^19^ Participants provide biospecimens, consent to research access to their VA health records, and complete surveys about family health history, health behaviors, military and environmental exposures, and other health-related factors. For the present analyses, we used data from 585,418 male MVP participants to develop and cross-validate the P-CARE model. All study participants provided blood samples for DNA extraction and genotyping using a custom Affymetrix Axiom biobank array containing 723,305 variants, enriched for low-frequency variants in African and Hispanic populations. Details on genotyping quality control and imputation have been described previously.^22^ Family history was defined as the presence or absence of one or more first-degree relatives with prostate cancer, as reported on the MVP survey. As described previously,^18,23^ prostate cancer diagnosis, age at diagnosis, and date of last follow-up were retrieved from the VA Corporate Data Warehouse based on International Classification of Diseases (ICD) diagnosis codes and VA Central Cancer Registry data. Age at diagnosis of metastasis (nodal and/or distant, regardless of whether metastases were detected at diagnosis or at recurrence) was determined via a validated natural language processing tool developed in the VA system.^18,24^ Cause and date of death were obtained from the National Death Index. Fatal prostate cancer was defined by ICD9 code 185 or ICD10 code C61 as underlying cause of death.

### PRACTICAL Consortium

Data from 4 external cohorts from the Prostate Cancer Association Group to Investigate Cancer Associated Alterations in the Genome (PRACTICAL) Consortium were used to externally validate the P-CARE model. Data from 18,457 men previously genotyped via OncoArray or iCOGs arrays^25,26^ were split into four datasets described in prior studies evaluating polygenic scores: 1) men of African ancestry (*n*=6,253); 2) men of Asian ancestry (*n*=2,320); 3) the Cohort of Swedish Men (COSM) population-based cohort with long-term outcomes (n=3,415); and 4) the population-based Prostate Testing for Cancer and Treatment (ProtecT) screening trial (*n*=6,411).^20^ Family history was defined as the presence or absence of a first-degree relative with a prostate cancer diagnosis. Clinically significant prostate cancer was defined previously as any case with Gleason score ≥7, PSA ≥10 ng/mL, T3-T4 stage, nodal metastases, or distant metastases.^20^ The COSM dataset additionally had age at prostate cancer death,^27^ and the ProtecT dataset had prostate biopsy results for both cases and controls with screening PSA ≥3 ng/mL.^28,29^

### Candidate variants and training for polygenic score

We considered variants previously identified from the following sources for potential inclusion in an updated polygenic score for the P-CARE model: 290 variants from a prior score, 613 variants identified as prostate cancer susceptibility loci in a multi-ancestry genome-wide association studies, 23 variants identified as susceptibility loci for benign elevation of prostate-specific antigen (PSA) or benign prostatic hypertrophy (BPH), 9 variants identified as prostate cancer susceptibility loci in men of African ancestry in a genome-wide meta-analysis, and 128 variants identified as susceptibility loci for prostate cancer in a genome-wide multi-ancestry meta-analysis.^20,30–33^ A machine-learning, least absolute shrinkage and selection operator (LASSO)-regularized Cox proportional hazards model approach was used in the MVP dataset to select the final variants for the polygenic score and estimate weights, using the R (v.4.4) “glmnet” package (v.4.1.8), as described previously.^34–36^ To develop the polygenic score, age at any prostate cancer diagnosis in MVP was the time to event, as this gives the most statistical power; controls were censored at age of last follow-up. First, we identified pairs of variants with highly correlated genotype (defined as *r*^2^ > 0.95) and used univariable Cox models to exclude the variant from each pair with weakest univariable association. Next, all remaining candidate variants were evaluated for inclusion in the new polygenic score using a Cox model with genotype allele counts of candidate variants and the first five FastPop principal components as predictor variables. Genetic principal components were estimated using 2,309 ancestry informative markers from FastPop.^37^ Loadings for the first 5 principal components were estimated in the 1000 Genomes Phase 3 dataset.^38^ The final form of the LASSO model was estimated using the lambda value that minimized the mean cross-validated error.^39^

We then used Cox proportional hazards models to evaluate the association of the new polygenic score with age at diagnosis of prostate cancer, age at diagnosis of nodal and/or distant metastatic prostate cancer, and age at prostate cancer death within the MVP dataset overall and in analyses stratified by continental population ancestry group, as described previously.^18,40^ Similarly, Cox models were used to evaluate the association between the new score and age at diagnosis of any prostate cancer, clinically significant prostate cancer, and fatal prostate cancer (in the COSM dataset) in the PRACTICAL cohort, as previously described.^18^

### P-CARE model development and validation

The resulting polygenic score was then carried forward for use in the development of an integrated clinical prediction model within MVP. We developed a Cox model for age at prostate cancer diagnosis as a function of the polygenic score, modeled as a continuous variable; family history of prostate cancer, modeled as a binary variable indicating presence or absence of at least one first-degree relative with prostate cancer; and population structure, modeled using the first two genetic principal components (PCs). Prior analyses showed that the first two PCs are sufficient to capture genetic variation for prostate cancer risk stratification compared to 5-10 PCs.^41^ Individuals not meeting the endpoint of interest were censored at last follow-up.

The resulting P-CARE model was then validated internally within the MVP dataset and externally within the 4 PRACTICAL datasets. Where available, we evaluated the association of the P-CARE model with age of diagnosis of any prostate cancer, clinically significant prostate cancer, metastatic prostate cancer, and fatal prostate cancer. As in our prior work,^20,27,34,41–46^ we estimated effect sizes using hazard ratios (HRs) and 10 iterations of 10-fold cross validation, calculated to make the following comparisons: HR_80/20_, men in the highest 20% versus lowest 20%; HR_95/50_, men in the highest 5% versus those with median values; and HR_20/50_, men in the lowest 20% vs those with median values. Within the MVP dataset, we generated cumulative incidence curves for each prostate cancer endpoint by P-CARE percentile groups, as previously described.^34,44^ We additionally generated cumulative incidence curves by P-CARE risk categories defined by risk of metastatic disease, given its morbidity and mortality and to counter the criticism that current prostate cancer screening approaches over-detect indolent disease.^12–15^ The high risk category was defined as an overall P-CARE HR>1.5 for metastatic prostate cancer and the low risk category was defined as HR<0.75 (consistent with routine clinical prediction tools for other diseases, such as breast cancer, diabetes, and cardiovascular disease^47–49^); all other risk values were considered average risk. Because ProtecT systematically collected prostate biopsies, this dataset offered the opportunity to correlate PSA values with likelihood of clinically significant prostate cancer. Within the ProtecT dataset, we calculated the positive predictive value (PPV) of a PSA value ≥3 ng/mL for clinically significant prostate cancer on biopsy among participants in the top 5th (PPV_95_) and top 20th P-CARE percentile (PPV_80_).^20,29^

### Clinical laboratory assay development and validation

The P-CARE model was then carried forward to develop a clinical laboratory assay (Broad Clinical Labs, Burlington, MA, USA) to enable precision prostate cancer screening informed by both the model and relevant rare variants, given their importance in prostate cancer risk.

### Blended genome exome assay

We constructed the assay on a novel, cost-efficient blended genome exome (BGE) platform, which combines 2-3x whole genome sequencing (WGS) with 60-90x exome sequencing in a single sequenced sample and has achieved >99% concordance with 30x genome sequencing data for both exome and genome short variants.^50^ Short variant calling was performed over the high coverage exome target regions using the Illumina DRAGEN Bio-IT platform version 4.2.7. Genotypes and dosage information over the whole genome were obtained from sequencing data through GLIMPSE2 imputation^51^ using the gnomAD HGDP and 1000 Genomes callset.^52^ Copy number variation was detected over the exome target regions using GATK-gCNV.^53^

### Analytic and clinical laboratory validation of polygenic score and P-CARE model

The analytic validity of the BGE platform for the polygenic score was assessed by comparing 60 clinical samples with previously-identified variants; reference samples from Coriell Institute for Medical Research with curated reference variant data sets maintained by the National Institute of Standards and Technology; and samples with known SNVs, indels, and CNVs from a combination of previous in-production clinical samples, previous eMERGE studies, previous CAP proficiency testing samples, Coriell samples, and the Coriell Ancestral Panel. For each of these samples, representing 6 genetic ancestry groups (Admixed American, African, Non-Finnish European, East Asian, South-East Asian, Ashkenazi), we generated both BGE and WGS data and calculated the PHS and genetic principal components. Additional evidence of clinical validity for both the polygenic score and the P-CARE model was obtained using 74,331 samples from the All of Us (AoU) Research Program with short-read WGS data and male sex assigned at birth, excluding samples flagged for failing QC criteria, for being related, or for lack of available electronic health record data. Polygenic score and genetic principal components were calculated from the WGS genotypes provided by AoU. Individuals were classified as cases and controls based on the presence or absence of “Malignant tumor of prostate” entries in the AoU electronic health record data. P-CARE values were calculated for each AoU participant using polygenic score, the first two genetic principal components, first-degree family history of prostate cancer, and the MVP-derived coefficients. To determine the association between P-CARE and prostate cancer case status in AoU, we calculated odds ratios for an individual to be diagnosed with prostate cancer in the low and high P-CARE categories, relative to the average category, using logistic regression models controlling for age.

### Rare variant selection, validation, and interpretation

We identified known prostate cancer-associated genes for which the National Comprehensive Cancer Network has issued clinical management recommendations.^54–56^ This gene list informed the filtering for an *in silico* gene panel for rare variant analysis. The ability of the BGE to identify pathogenic or likely pathogenic variants in these genes was evaluated by assessing the overall technical performance of 12 genes related to hereditary prostate cancer risk (*BRCA1, BRCA2, ATM, PALB2, CHEK2, HOXB13, MLH1, MSH2, MSH6, PMS2, TP53*, and *EPCAM*) and identification of known variants from previous clinical testing (SNVs, small InDels, CNVs) within these genes in 18 clinical samples. Technical performance of these genes was assessed by determining the percentage of undercovered bases within a panel gene. A base is considered covered if it satisfies the following: coverage >20X, base quality >20, and mapping quality >20. This coverage analysis was performed with two sample fraction thresholds: ≥80% and ≥20%.

We determined the sensitivity for the detection of rare monogenic variants if the variant of interest was identified in the variant call file and would meet quality and prioritization metrics to be flagged for manual review by our tertiary analysis platform. Additionally, inter and intra run precision was assessed by running samples in triplicate across different runs and within the same run, respectively. We developed a workflow to classify, review, and prioritize variants in a tertiary analysis platform (Fabric Genomics, Oakland, CA, USA) prior to in-house clinical interpretation and reporting of pathogenic and likely pathogenic variants by a team of board-certified geneticists.

### Clinical report development

After clinical laboratory validation of the P-CARE and rare variant pipelines, we developed a laboratory report package suitable for the clinical implementation of these results, consistent in format and content with other clinical genetic test reports and with our prior work.^4,5,57^ As described in the Results, the report package consisted of separate laboratory reports for the

P-CARE and rare variant results and a summary report synthesizing the result types and providing prostate cancer screening recommendations for the patient and provider.

## RESULTS

### Association of polygenic score with prostate cancer outcomes

The final model included 601 of the 707 unique candidate variants evaluated (**Supplemental File 1**). The resulting polygenic hazard score (PHS_601_) was associated with age at diagnosis of prostate cancer, metastatic prostate cancer, and prostate cancer death in MVP (**Table 1**).

**Table 1:** Association of polygenic score with prostate cancer outcomes in MVP and PRACTICAL cohorts. Association of PHS_601_ with any, metastatic, and fatal prostate cancer in MVP (total and genetic ancestry-stratified groups) and with any, clinically significant, and fatal prostate cancer in four PRACTICAL consortium datasets. Abbreviations: CI, confidence interval; COSM, Cohort of Swedish Men; HR, hazard ratio; MVP, Million Veteran Program; PHS, polygenic hazard score; PRACTICAL, Prostate Cancer Association Group to Investigate Cancer Associated Alterations in the Genome; ProtecT, Prostate Testing for Cancer and Treatment.

**Figure 3: Positive predictive value of PSA in ProtecT by P-CARE values.**
| Clinical endpoint | N | HR (95% CI) |  |  |  |  |
| --- | --- | --- | --- | --- | --- | --- |
|  |  | HR<sub>SD</sub> | HR<sub>80/20</sub> | HR<sub>20/50</sub> | HR<sub>80/50</sub> | HR<sub>95/50</sub> |
| MVP development and validation |  |  |  |  |  |  |
| Any prostate cancer |  |  |  |  |  |  |
| All | 585,418 | 2.02 (1.97 - 2.07) | 6.27 (5.85 - 6.72) | 0.43 (0.41 - 0.45) | 2.72 (2.62 - 2.82) | 4.06 (3.86 - 4.29) |
| African | 105,014 | 1.94 (1.84 - 2.06) | 5.81 (5.06 - 6.79) | 0.48 (0.45 - 0.52) | 2.81 (2.59 - 3.08) | 3.74 (3.37 - 4.23) |
| European | 420,722 | 2.04 (1.97 - 2.11) | 5.74 (5.26 - 6.20) | 0.43 (0.41 - 0.45) | 2.46 (2.35 - 2.55) | 3.78 (3.54 - 4.03) |
| American | 50,590 | 2.03 (1.83 - 2.26) | 5.55 (4.24 - 7.07) | 0.44 (0.39 - 0.50) | 2.44 (2.13 - 2.79) | 3.72 (3.06 - 4.52) |
| East Asian | 9,092 | 2.10 (1.59 - 2.81) | 5.81 (2.90 - 10.65) | 0.44 (0.32 - 0.60) | 2.41 (1.72 - 3.36) | 3.83 (2.31 - 6.04) |
| Metastatic prostate cancer |  |  |  |  |  |  |
| All | 585,418 | 2.07 (1.95 - 2.17) | 6.69 (5.70 - 7.62) | 0.42 (0.40 - 0.45) | 2.81 (2.58 - 3.01) | 4.27 (3.78 - 4.71) |
| African | 105,014 | 1.90 (1.65 - 2.19) | 5.60 (3.81 - 7.84) | 0.50 (0.43 - 0.58) | 2.73 (2.18 - 3.39) | 3.62 (2.70 - 4.77) |
| European | 420,722 | 1.96 (1.80 - 2.19) | 5.20 (4.19 - 6.83) | 0.45 (0.39 - 0.50) | 2.33 (2.08 - 2.68) | 3.50 (2.97 - 4.29) |
| American | 50,590 | 2.07 (1.42 - 2.64) | 6.02 (2.33 - 10.47) | 0.44 (0.32 - 0.67) | 2.51 (1.55 - 3.40) | 3.92 (1.91 - 6.08) |
| East Asian | 9,092 | 3.11 (0.59 - 8.64) | 63.26 (0.31 - 154.75) | 0.51 (0.08 - 1.77) | 4.19 (0.56 - 12.63) | 16.90 (0.40 - 44.15) |
| Fatal prostate cancer |  |  |  |  |  |  |
| All | 585,418 | 1.96 (1.75 - 2.18) | 5.81 (4.31 - 7.65) | 0.45 (0.40 - 0.51) | 2.60 (2.22 - 3.03) | 3.82 (3.06 - 4.72) |
| African | 105,014 | 1.62 (1.16 - 2.12) | 3.81 (1.49 - 7.26) | 0.61 (0.44 - 0.85) | 2.15 (1.27 - 3.22) | 2.68 (1.35 - 4.45) |
| European | 420,722 | 1.92 (1.60 - 2.20) | 4.99 (3.18 - 6.86) | 0.47 (0.39 - 0.57) | 2.27 (1.81 - 2.69) | 3.39 (2.41 - 4.32) |
| American | 50,590 | 2.22 (0.98 - 4.25) | 8.45 (0.95 - 32.23) | 0.48 (0.19 - 1.03) | 2.78 (0.97 - 6.14) | 4.81 (0.96 - 14.32) |
| East Asian | 9,092 | 7.97E56 (0.93 - 9.39E40) | 1.56E34 (0.88 - 5.4E109) | 3.27E112 (6.85E-47 - 1.07) | 1.28E70 (0.93 - 1.82E56) | 3.04E108 (0.90 - 7.71E83) |
| PRACTICAL replication |  |  |  |  |  |  |
| Any prostate cancer |  |  |  |  |  |  |
| COSM | 3,415 | 2.27 (2.11 - 2.46) | 9.18 (6.66 - 12.93) | 0.36 (0.31 - 0.42) | 3.34 (2.97 - 3.98) | 5.35 (4.26 - 6.92) |
| ProtecT | 6,411 | 1.87 (1.78 - 2.01) | 5.67 (4.75 - 6.73) | 0.44 (0.40 - 0.48) | 2.78 (2.47- 3.05) | 3.78 (3.31 - 4.29) |
| African | 6,253 | 1.84 (1.72 - 1.98) | 8.55 (6.64 - 11.08) | 0.41 (0.36 - 0.46) | 3.47 (2.93 - 3.99) | 4.49 (3.71 - 5.37) |
| Asian | 2,320 | 2.15 (1.92 - 2.38) | 8.80 (6.73 - 11.09) | 0.35 (0.30 - 0.39) | 3.02 (2.63 - 3.39) | 5.26 (4.24 - 6.48) |
| Clinically significant prostate cancer |  |  |  |  |  |  |
| COSM | 3,415 | 2.30 (2.09 - 2.53) | 9.35 (7.32 - 12.20) | 0.36 (0.31 - 0.41) | 2.81 (2.58 - 3.01) | 4.27 (3.78 - 4.71) |
| ProtecT | 6,411 | 2.02 (1.88 - 2.21) | 7.02 (5.65 - 8.42) | 0.40 (0.36 - 0.44) | 2.73 (2.18 - 3.39) | 4.45 (3.76 - 5.11) |
| African | 6,253 | 1.85 (1.69 - 2.02) | 8.61 (6.50 - 11.61) | 0.41 (0.35 - 0.47) | 2.51 (1.55 - 3.40) | 3.92 (1.91 - 6.08) |
| Asian | 2,320 | 2.11 (1.90 - 2.39) | 7.88 (5.94 - 10.68) | 0.36 (0.32 - 0.42) | 2.85 (2.44 - 3.32) | 4.83 (3.84 - 6.09) |
| Fatal prostate cancer |  |  |  |  |  |  |
| COSM | 3,415 | 1.91 (1.65 - 2.28) | 5.88 (3.51 - 9.19) | 0.45 (0.36 - 0.56) | 2.58 (1.97 - 3.32) | 3.82 (2.59 - 5.35) |

Among the overall MVP cohort, the HR per standard deviation increase in PHS_601_ for prostate cancer, metastatic prostate cancer, and prostate cancer death were 2.02 (95% CI 1.97-2.07),

2.07 (95% CI 1.95-2.17), and 1.96 (95% CI 1.75-2.18), respectively. The associations between PHS_601_ and prostate cancer outcomes were similar in each ancestry-stratified analysis with >100 events in MVP and within each ancestry-specific PRACTICAL dataset (**Table 1**). Among the American and East Asian subgroups in MVP, which had small case numbers, associations with metastatic and fatal prostate cancer were not statistically significant but had consistent directions of effect.

### Association of P-CARE with prostate cancer outcomes

Similarly, the P-CARE score that integrated PHS_601_, genetic ancestry, and family history described a strong gradient of risk for any, clinically significant, metastatic, and fatal prostate cancer across MVP and PRACTICAL datasets (**Table 2**). Among the overall MVP cohort, the HR per standard deviation increase in P-CARE for prostate cancer, metastatic prostate cancer, and prostate cancer death were 2.04 (95% CI 1.99-2.08), 2.05 (95% CI 1.93-2.16), and 1.95 (95% CI 1.76-2.15), respectively. Across the MVP and PRACTICAL datasets, compared to men with median P-CARE values, men in the lowest P-CARE quintile had HR 0.39-0.45 for the 4 prostate cancer outcomes (HR_20/50_), while men in the highest P-CARE quintile had HR 2.75-4.03 (HR_80/50_, **Table 2**). The direction and magnitude of association between P-CARE and the prostate cancer outcomes were similar in analyses of subgroups defined by genetic ancestry (**Supplemental Table 5**) and, alternatively, by self-reported race and ethnicity (**Supplemental Table 6**), in each subgroup with adequate case counts. Within the ProtecT dataset, the PPV of a PSA value ≥3 ng/mL for clinically significant prostate cancer was 0.13 (95% CI 0.12-0.14) in the overall dataset and 0.19 (95% CI 0.16-0.21) and 0.23 (0.17-0.28) in the subsets in the top 20% and top 5% of P-CARE values, respectively (Figure 3).

**Table 2:**
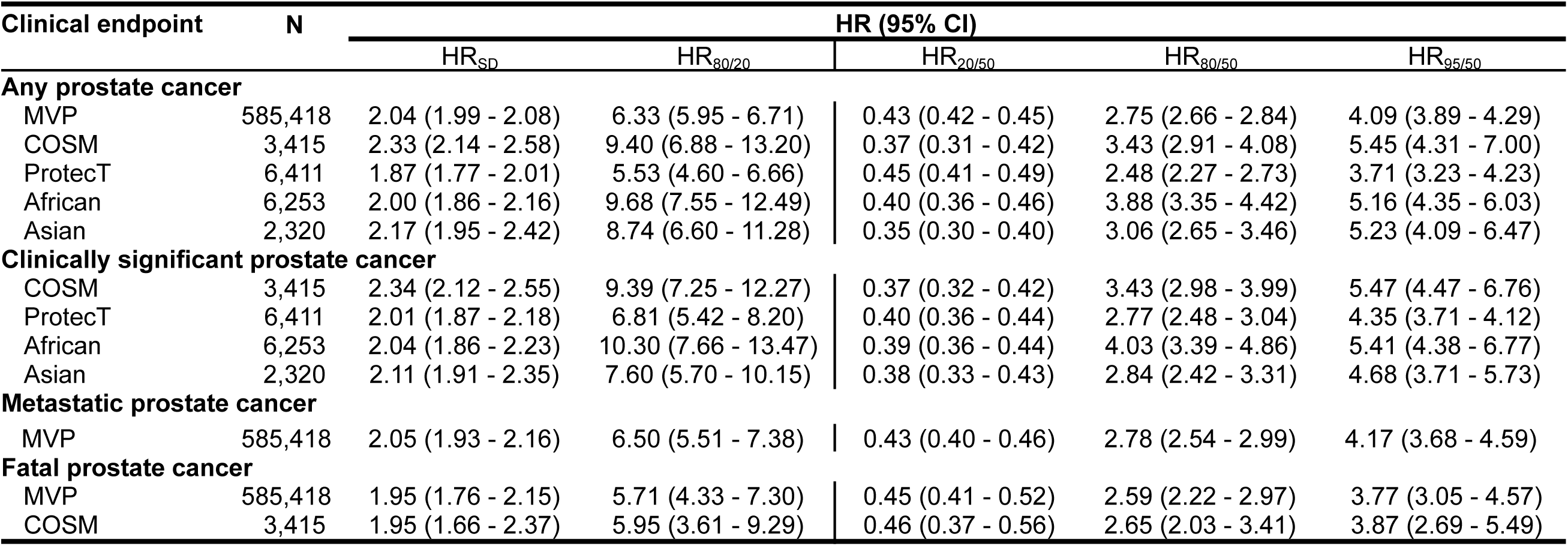
Association of P-CARE model with prostate cancer outcomes in MVP and PRACTICAL cohorts. Association of P-CARE model with any prostate cancer, clinically significant prostate cancer, metastatic prostate cancer, and fatal prostate cancer in MVP and four PRACTICAL consortium datasets. As described, P-CARE model consists of PHS_601_, first-degree family history of prostate cancer, and genetic principal components. Abbreviations: CI, confidence interval; COSM, Cohort of Swedish Men; HR, hazard ratio; MVP, Million Veteran Program; P-CARE, Prostate CA Risk and Evaluation; PRACTICAL, Prostate Cancer Association Group to Investigate Cancer Associated Alterations in the Genome; ProtecT, Prostate Testing for Cancer and Treatment.

| Clinical endpoint | N | HR (95% CI) |  |  |  |  |
| --- | --- | --- | --- | --- | --- | --- |
|  |  | HR<sub>SD</sub> | HR<sub>80/20</sub> | HR<sub>20/50</sub> | HR<sub>80/50</sub> | HR<sub>95/50</sub> |
| Any prostate cancer |  |  |  |  |  |  |
| MVP | 585,418 | 2.04 (1.99 - 2.08) | 6.33 (5.95 - 6.71) | 0.43 (0.42 - 0.45) | 2.75 (2.66 - 2.84) | 4.09 (3.89 - 4.29) |
| COSM | 3,415 | 2.33 (2.14 - 2.58) | 9.40 (6.88 - 13.20) | 0.37 (0.31 - 0.42) | 3.43 (2.91 - 4.08) | 5.45 (4.31 - 7.00) |
| ProtecT | 6,411 | 1.87 (1.77 - 2.01) | 5.53 (4.60 - 6.66) | 0.45 (0.41 - 0.49) | 2.48 (2.27 - 2.73) | 3.71 (3.23 - 4.23) |
| African | 6,253 | 2.00 (1.86 - 2.16) | 9.68 (7.55 - 12.49) | 0.40 (0.36 - 0.46) | 3.88 (3.35 - 4.42) | 5.16 (4.35 - 6.03) |
| Asian | 2,320 | 2.17 (1.95 - 2.42) | 8.74 (6.60 - 11.28) | 0.35 (0.30 - 0.40) | 3.06 (2.65 - 3.46) | 5.23 (4.09 - 6.47) |
| Clinically significant prostate cancer |  |  |  |  |  |  |
| COSM | 3,415 | 2.34 (2.12 - 2.55) | 9.39 (7.25 - 12.27) | 0.37 (0.32 - 0.42) | 3.43 (2.98 - 3.99) | 5.47 (4.47 - 6.76) |
| ProtecT | 6,411 | 2.01 (1.87 - 2.18) | 6.81 (5.42 - 8.20) | 0.40 (0.36 - 0.44) | 2.77 (2.48 - 3.04) | 4.35 (3.71 - 4.12) |
| African | 6,253 | 2.04 (1.86 - 2.23) | 10.30 (7.66 - 13.47) | 0.39 (0.36 - 0.44) | 4.03 (3.39 - 4.86) | 5.41 (4.38 - 6.77) |
| Asian | 2,320 | 2.11 (1.91 - 2.35) | 7.60 (5.70 - 10.15) | 0.38 (0.33 - 0.43) | 2.84 (2.42 - 3.31) | 4.68 (3.71 - 5.73) |
| Metastatic prostate cancer |  |  |  |  |  |  |
| MVP | 585,418 | 2.05 (1.93 - 2.16) | 6.50 (5.51 - 7.38) | 0.43 (0.40 - 0.46) | 2.78 (2.54 - 2.99) | 4.17 (3.68 - 4.59) |
| Fatal prostate cancer |  |  |  |  |  |  |
| MVP | 585,418 | 1.95 (1.76 - 2.15) | 5.71 (4.33 - 7.30) | 0.45 (0.41 - 0.52) | 2.59 (2.22 - 2.97) | 3.77 (3.05 - 4.57) |
| COSM | 3,415 | 1.95 (1.66 - 2.37) | 5.95 (3.61 - 9.29) | 0.46 (0.37 - 0.56) | 2.65 (2.03 - 3.41) | 3.87 (2.69 - 5.49) |

P-CARE risk categories defined by thresholds of HR 0.75 and HR 1.5 for metastatic prostate cancer are shown in **Table 3**. Overall, the model categorized 25.1%, 37.3%, and 37.6% of MVP participants as low-, average-, and high-risk, respectively. The model categorized 68.7% of participants with positive family history as high-risk and only 5.6% as low-risk. Among participants self-reporting Black or African-American race, only 2.8% were categorized as low risk. Figure 2 shows cumulative prostate cancer incidence curves in MVP both by P-CARE percentile groups and by P-CARE risk category. As shown in **Table 4**, by age 80, men in the high-risk P-CARE group had a cumulative risk of any, metastatic, and fatal prostate cancer of 37.4%, 4.4%, and 0.8%, respectively.

**Figure 2:**
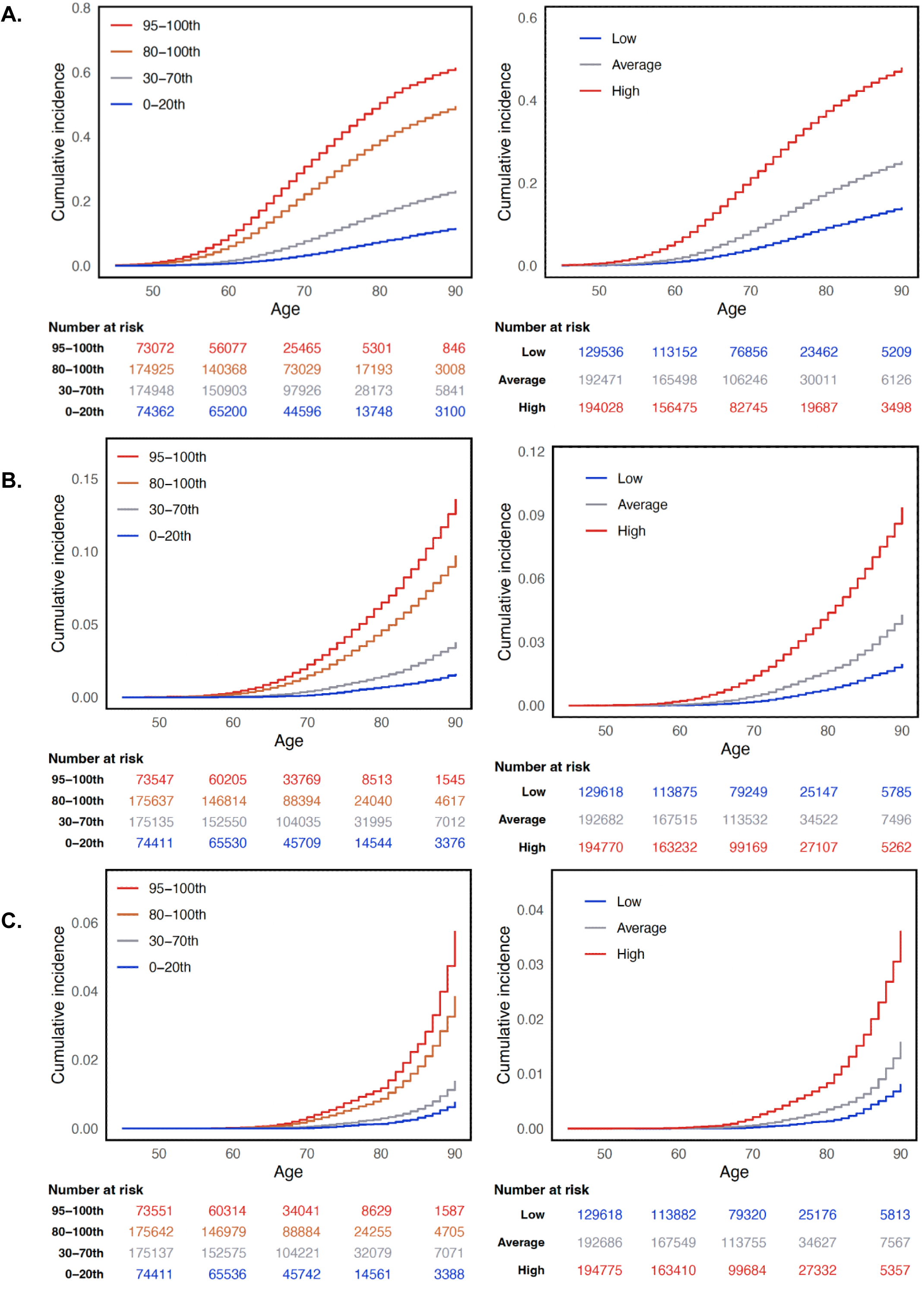
Prostate cancer cause-specific cumulative incidence in MVP by P-CARE strata. Cause specific cumulative incidence within MVP for (A) prostate cancer, (B) metastatic prostate cancer, and (C) fatal prostate cancer. The left column shows incidence for each endpoint by P-CARE percentile group: 0-20^th^, 30-70^th^, 80-100^th^, and 95-100^th^. The right column shows incidence for each endpoint by P-CARE risk category: high, average, and low risk. Abbreviations: MVP, Million Veteran Program; P-CARE, Prostate CA Risk and Evaluation.

**Figure 3.**
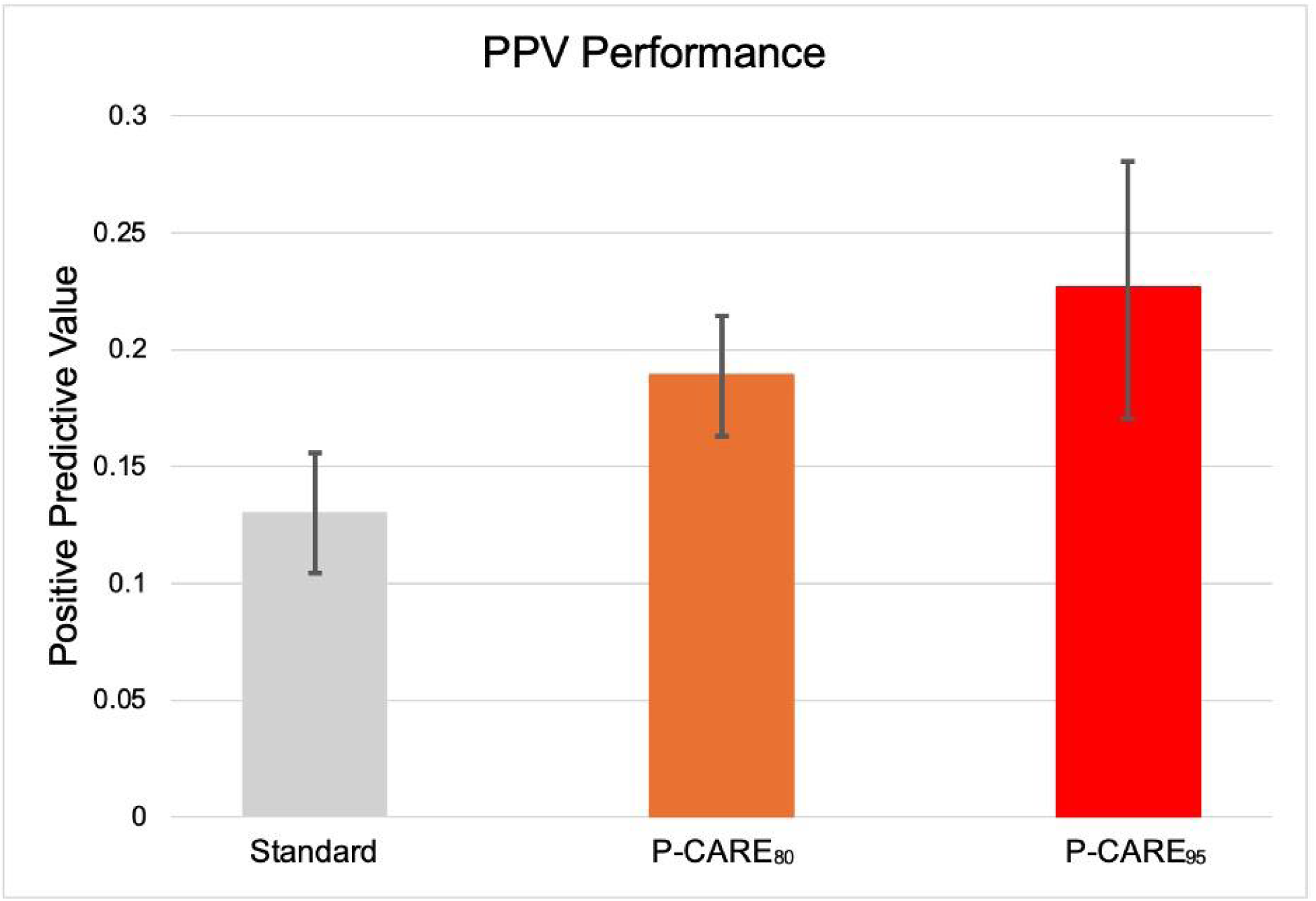
Positive predictive value of PSA in ProtecT by P-CARE values. Illustrated are PPV (95% CI) for a PSA value ≥3 ng/mL for clinically significant prostate cancer among three groups of men in the ProtecT study: all men (regardless of P-CARE value), men in the top 20% of P-CARE values (P-CARE_80_), and men in the top 5% of P-CARE values (P-CARE_95_). Abbreviations: CI, confidence interval; P-CARE, Prostate CA Risk and Evaluation; PPV, positive predictive value; ProtecT, Prostate Testing for Cancer and Treatment; PSA, prostate-specific antigen.

**Table 3:** Characteristics of P-CARE risk categories among 585,418 MVP participants. MVP, Million Veteran Program; P-CARE, Prostate CA Risk and Evaluation.

|  | N | P-CARE risk category, N (%) |  |  |
| --- | --- | --- | --- | --- |
|  |  | Low risk (HR<0.75) | Average risk (HR 0.75-1.5) | High risk (HR>1.5) |
| **Total** | 585,418 | 146,826 (25.08) | 218,530 (37.33) | 220,062 (37.59) |
| **Positive family history** | 28,358 | 1,595 (5.62) | 7,270 (25.63) | 19,493 (68.73) |
| **Genetic ancestry groups** |  |  |  |  |
| African | 105,014 | 2,607 (2.48) | 19,314 (18.39) | 83,093 (79.12) |
| European | 420,722 | 128,096 (30.44) | 173,774 (41.30) | 118,852 (28.24) |
| American | 50,590 | 12,842 (25.38) | 21,518 (42.53) | 16,230 (32.08) |
| East Asian | 9,092 | 3,281 (36.08) | 3,924 (43.15) | 1,887 (20.75) |
| **Self-reported race/ethnicity groups** |  |  |  |  |
| American Indian or Alaska Native | 5507 | 1,346 (24.44) | 2,236 (40.60) | 1,925 (34.95) |
| Asian | 6210 | 2,403 (38.69) | 2,684 (43.22) | 1,123 (18.08) |
| Black or African American | 101,920 | 2,812 (2.75) | 18,986 (18.62) | 80,122 (78.61) |
| Hispanic White | 26,037 | 6,871 (26.38) | 11,067 (42.50) | 8,099 (31.10) |
| Native Hawaiian or Pacific Islander | 3042 | 755 (24.81) | 1,259 (41.38) | 1,028 (33.79) |
| Non-Hispanic White | 418,387 | 126,633 (30.26) | 172,661 (41.26) | 119,093 (28.46) |
| Other | 8077 | 1,938 (23.99) | 3,303 (40.89) | 2,836 (35.11) |
| Unknown | 16,238 | 4,068 (25.05) | 6,334 (39.00) | 5,836 (35.94) |

**Table 4:**
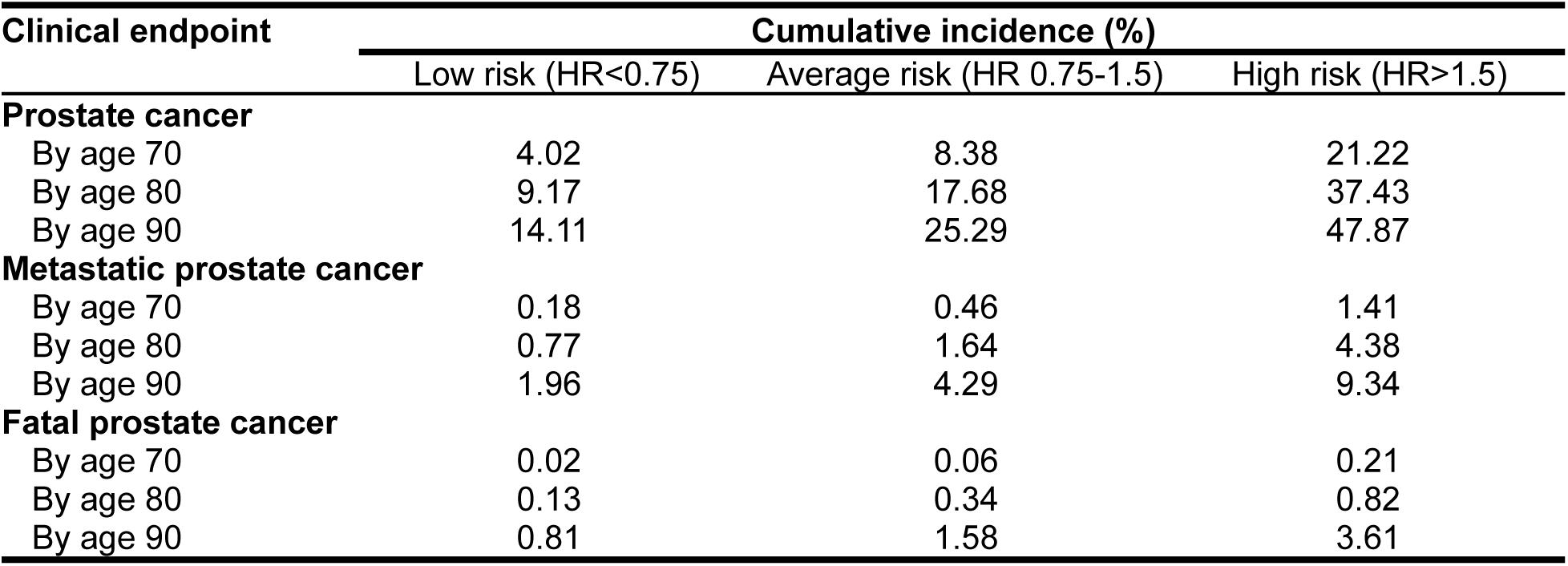
Prostate cancer cause-specific cumulative incidence in MVP by P-CARE category. Abbreviations: HR, hazard ratio; MVP, Million Veteran Program; P-CARE, Prostate CA Risk and Evaluation.

### Clinical laboratory assay validation

Compared to WGS data, scores derived from the BGE platform achieved a Pearson correlation of r > 0.998 for PHS_601_ and r > 0.999 for both principal components. For the 12 genes related to hereditary prostate cancer risk, 11 out of the 12 genes met thresholds for coverage and callability. The one gene that did not pass technical assessment was *PMS2*, which is well known as a challenging gene due to a highly homologous pseudogene. Within the *PMS2* gene, exon 13, 14 and exons 13, 14, 15 were undercovered for sample fractions of 0.80 and 0.20, respectively. Of the 18 samples assessed for monogenic rare variants 18/18 variants of interest were identified in the variant call file (7/7 SNVs, 5/5 InDels and 6/6 CNVs). However, copy number variant calls that fall below certain quality thresholds will be filtered out in the interpretation process. These thresholds are defined as: QUAL ≥ 50 for duplications, QUAL ≥ 100 for heterozygous deletions, and QUAL ≥ 400 for homozygous deletions. Three out of the six copy number variant calls were of low quality and would not have been clinically reported. This finding is consistent with assay limitations for small copy number variants of less than three exons in size showing reduced sensitivity as reported in the technical validation. Precision for variant of interest detection was seen at 100% for intra and inter run concordance.

Within the AoU dataset, the PHS_601_ was associated with prostate cancer with an odds ratio per standard deviation of 1.91 (95% CI: 1.85-1.98). In the same dataset, for the full P-CARE model (PHS_601_ plus genetic principal components and family history) we found an odds ratio of 2.41 (95% CI: 2.25-2.60) for individuals in the high-risk category to be diagnosed with prostate cancer, compared to individuals classified as average risk. Similarly, individuals in the low-risk category show an odds ratio of 0.48 (95% CI: 0.44-0.54), compared to individuals classified as average risk. Notably, this strong association holds across different ancestries. (**Supplemental Figure 1**)

### Clinical P-CARE and monogenic reports

An example of the resulting laboratory report package is shown in **Supplemental File 2**. The cover page summarizes the results of both the monogenic and P-CARE analyses and provides an overall risk category for the individual based on these results. An individual with a pathogenic or likely pathogenic variant in one of the 12 prostate cancer-associated genes is categorized as high-risk, regardless of P-CARE results. Individuals without such a variant are categorized as low-, average-, or high-risk according to their P-CARE result, with thresholds at HR=0.75 and HR=1.5, as described in the Methods. The cover page also links these risk categories to tailored prostate cancer screening recommendations for the individual. After this cover page summary, separate P-CARE and rare variant reports provide further detail about these individual result types, including information about P-CARE model development and validation, technical descriptions of the analyses performed, relevant gene and disease information, and literature references. These reports are now being used in the national ProGRESS randomized clinical trial, in which 5,000 prostate cancer screen-eligible VA patients are randomly assigned to usual care versus precision screening recommendations informed by P-CARE and rare variants.

## DISCUSSION

We used genomic, clinical, and survey data from a large national biobank to develop a genomics-informed prostate cancer prediction model consisting of family history, genetic principal components, and an updated polygenic score of 601 prostate trait-associated loci. Patients in the lowest and highest 20% of values under this model have 0.4-fold and 2.7-fold risk of prostate cancer, respectively, compared to those with median values; replication in external multiancestry cohorts confirmed these associations. Men at highest risk of developing advanced prostate cancer are most likely to benefit from screening; the P-CARE model is associated with risk of all, clinically significant, metastatic, and fatal prostate cancer. When low and high risk were defined as HR<0.75 and HR>1.5, respectively, for metastatic prostate cancer, the cumulative incidence of metastatic prostate cancer by age 80 in the biobank was 0.8% in the low-risk group and 4.4% in the high-risk group. We then developed and validated a clinical assay on a cost-efficient BGE platform for both the prediction model and rare pathogenic variants in known prostate cancer genes. This assay and associated clinical reports are now enabling a clinical trial of precision prostate cancer screening among patients receiving care from the national healthcare system from which the biobank data were derived. This approach illustrates the power of genomics-enabled learning health systems to generate translatable discoveries for implementation in preventive healthcare.

We designed the P-CARE model and ongoing prostate cancer screening trial to examine how the routine collection and interpretation of genomic data in preventive care might improve upon existing screening practices in a large integrated health system. Prostate cancer is highly prevalent, but despite randomized controlled trial evidence that screening with PSA testing can reduce prostate cancer mortality,^14,58^ guidelines vary by organization and country^11^ on how to balance the benefits of screening (early detection and treatment, resulting in lower incidence of advanced and lethal disease) and its potential risks (overdiagnosis of apparently indolent disease and morbidity from unnecessary procedures and treatments). As a result, screening practices are highly variable.^16,17,59–62^ Better models are needed to distinguish men most likely to benefit from screening from those for whom its risks might outweigh its benefits. A learning health system approach is ideal to improve prostate cancer screening for a few reasons. First, risk prediction models that inform the net benefit of cancer screening depend in large part on model calibration within a population; relative and absolute risk estimates derived from a healthcare system-linked biobank are thus particularly informative for patients receiving care in that system. In particular, age is a critical factor not only in the risk of advanced prostate cancer but also for the competing health risks that might make prostate cancer early detection less important;^63^ our time-to-event analysis and age-specific cumulative incidence curves account for age and allow physicians to balance these with age-related competing risks for a given individual to guide age-based screening decisions. Second, the effect sizes of polygenic scores themselves, including for prostate cancer, can vary between biobanks.^64,65^ Third, the net benefit of prostate cancer screening in a population is highly dependent on the downstream diagnostic and therapeutic management of elevated PSA values and abnormal prostate biopsy results;^66,67^ nesting the evaluation of a new screening paradigm within its target healthcare delivery system helps ensure that system-specific clinical practice patterns are included.

Our approach also seeks to address controversies in prostate cancer screening that are intimately intertwined with health disparities. In the United States, Black men are more likely to be both diagnosed with and die from prostate cancer.^68^ Possible causal factors include genetic, environmental, and social determinants of health, including systemic racism and access to screening and other healthcare.^69–71^ Black men are highlighted in prostate cancer guidelines as a group whose high risk merits earlier screening.^9,10^ This recommendation is appropriate to address racial disparities in prostate cancer outcomes. However, at the same time, the use of race in medical decision-making can inappropriately ascribe to biology effects that arise from a complex social construct confounded by myriad social determinants of health; it also ignores the complex multiracial and multiancestry backgrounds of individuals in modern healthcare system populations. We thus set out to develop a prostate cancer risk prediction model that did not include discrete race or genetic ancestry categories, favoring instead principal components as a continuous measure of genetic variation. At the same time, we confirmed that the resulting model performed well across categories of socially defined populations (race and ethnicity groups in MVP and external cohorts). Initial genome studies predominantly included individuals with European ancestry, but more recent work has improved genetic discovery and risk stratification in more diverse populations, including African ancestry.^20,30,32,41,43,46^ The P-CARE model extends this work, confirming that most Black men, but not all, have high risk. While the model does not fully disentangle the confounded associations between genetic ancestry and social determinants of prostate cancer risk, it represents an advance towards a more equitable, tailored approach to risk stratification and screening that does not treat race as a biological construct.

Family history of prostate cancer and certain rare genetic variants are also known prostate cancer risk factors, independent of ancestry and polygenic score.^20,41,43,46^ We designed the P-CARE model to build upon, not replace, these clinical risk factors, similar to breast cancer screening models.^72,73^ Rare variants in several genes, including *BRCA2* and *MSH2*, are known to increase prostate cancer risk and thus have separate screening guidelines for carriers.^54^ Carrier status of these variants is presently unknown for the vast majority of prostate cancer screen-eligible patients and yet might play a more prominent role in preventive care in a future when genomic testing is more commonplace. Despite aggregate analyses suggesting that polygenic scores can modify the effects of these rare variants,^74–76^ we determined that these modified associations are not yet robust enough for individual variant-level clinical reporting and should not supersede NCCN guidelines for the clinical management of rare variants. We therefore chose a genomic analysis platform that could detect and interpret these important rare variants and will report them according to established clinical guidelines to participants. By combining high-coverage exome and low-coverage whole genome sequencing data, the novel BGE technology provides a cost-effective, scalable and accurate platform for implementing the P-CARE model in clinical care. In the ProGRESS trial (ClinicalTrials.gov ID NCT05926102) participants and their healthcare providers are now receiving clinical reports with P-CARE results and the results of rare variant analysis, enabling an evaluation of a precision screening approach on prostate cancer in the U.S. Veterans Health Administration.

Our work has some limitations. Despite the strengths of our learning healthcare system approach described above, this system-specific model may not generalize to other settings with different population risks and screening practices. Model replication in the diverse PRACTICAL and All of Us datasets mitigates this concern, but other healthcare systems should examine model calibration in their own data before implementation. In addition, while the inclusion of family history, polygenic score, genetic principal components, and rare variants improves upon existing clinical prostate cancer screening approaches, the P-CARE model cannot disentangle the effects of genetic predisposition from environmental exposures and other social determinants that shape prostate cancer risk. Ongoing and future work should examine how to model and include other important risk factors in a clinically implementable risk stratification tool.^68^ Finally, BGE has many benefits including genome level variant information for PRS as well as an exome backbone for monogenic reporting, but there are limitations that come with an exome based approach that a purpose built capture panel may overcome, including lower sensitivity around complex regions of genes like *PMS2* and reduced sensitivity of small copy number variants below 3 exons in size.

In summary, a healthcare system-linked biobank has enabled the development, replication, and clinical laboratory validation of an updated prostate cancer risk model, now implemented in a clinical trial of precision prostate cancer screening. This approach exemplifies the power of genomics-enabled learning health systems to accelerate the discovery and translation of precision technologies to improve population health outcomes.

## Supporting information

Acknowledgements and Funding

## Data Availability

It is not possible for the authors to directly share the individual-level data that were obtained from the Million Veteran Program (MVP) due to constraints stipulated in the informed consent. Anyone wishing to gain access to this data should inquire directly to MVP at. The data generated from our analyses are included in the manuscript.

