## Supplement for "From a genomic risk model to clinical trial implementation in a learning health system: the ProGRESS Study"

**SUPPLEMENTAL FILES**

**Supplemental table 1: Description of Million Veteran Program (MVP) cohort**

**Supplemental table 2: Description of PRACTICAL consortium cohorts**

**Supplemental table 3: Association of family history with prostate cancer outcomes in MVP and PRACTICAL cohorts**

**Supplemental table 4: Coefficients in final P-CARE model**

**Supplemental table 5: Association of P-CARE model with prostate cancer outcomes in genetic ancestry strata in MVP**

**Supplemental table 6: Association of P-CARE model with prostate cancer outcomes in race- and ethnicity-stratified subgroups in MVP**

**Supplemental figure 1: Odds of prostate cancer in All of Us Research Program by P-CARE category**

**Supplemental file 1: Variants included in final $PHS_{601}$**

**Supplemental file 2: Template of laboratory report package for ProGRESS clinical trial**

| | Cohort | | | | |
|---|---|---|---|---|---|
| | All | African | European | American | East Asian |
| **Participants, number** | | | | | |
| All participants | 585,418 | 105,014 | 420,722 | 50,590 | 9,092 |
| Prostate cancer | 68,618 | 16,178 | 48,178 | 3775 | 487 |
| Metastases from prostate cancer | 6606 | 1726 | 4467 | 369 | 44 |
| Death from prostate cancer | 1709 | 364 | 1250 | 87 | 8 |
| **Age demographics, years** | | | | | |
| Age at enrollment, median (IQR) | 65 (56, 71) | 60 (52, 66) | 67 (59, 73) | 59 (45, 68) | 55 (40, 67) |
| Age at diagnosis, median (IQR) | 67 (62, 72) | 63 (58, 68) | 68 (63, 73) | 65 (60, 70) | 66 (61, 72) |
| Age at last follow-up, median (IQR) | 69 (60, 75) | 64 (57, 71) | 71 (63, 76) | 63 (49, 71) | 59 (43, 70) |

**Supplemental Table 1: Description of Million Veteran Program (MVP) cohort**. Ages and distribution of continental population ancestry groups among MVP participants included in analyses. Abbreviations: IQR, interquartile range.

| | Cohort | | | |
|---|---|---|---|---|
| | COSM | ProtecT | African | Asian |
| **Participants, number** | | | | |
| Total | 3279 | 6411 | 6253 | 2320 |
| Controls | 1116 | 4828 | 3013 | 1184 |
| Prostate cancer cases | 2163 | 1583 | 3240 | 1194 |
| Clinically significant prostate cancer cases[a] | 1403 | 628 | 1424 | 716 |
| Fatal prostate cancer cases | 278 | - | - | - |
| **Participant demographic information** | | | | |
| Age at diagnosis, median (IQR) | 70.0 (64.6, 76.5) | 63.4 (59.0, 67.0) | 62.8 (56.1, 68.1) | 68.2 (62.0, 74.0) |
| Age at last follow-up, median (IQR) | 78.1 (72.3, 84.1) | 60 (55.7, 64.4) | 63.0 (58, 72.1) | 72.9 (66.5, 78.0) |
| Positive family history, n | 355 | 394 | 999 | 94 |
| **P-CARE** | | | | |
| P-CARE score, median (IQR) | 20.36 (19.90, 20.83) | 20.16 (19.74, 20.59) | 21.04 (20.64, 21.44) | 20.10 (19.69, 20.51) |

**Supplemental Table 2: Description of PRACTICAL consortium cohorts.** Participant characteristics of the four PRACTICAL Consortium case-control cohorts used in external validation of P-CARE. [a]Clinically significant prostate cancer defined as Gleason score ≥7, PSA ≥10 ng/mL, T3-T4 stage, nodal metastases, or distant metastases. Abbreviations: CI, confidence interval; COSM, Cohort of Swedish Men; IQR, interquartile range; P-CARE, Prostate CA Risk and Evaluation; PRACTICAL, Prostate Cancer Association Group to Investigate Cancer Associated Alterations in the Genome; ProtecT, Prostate Testing for Cancer and Treatment; PSA, prostate-specific antigen.

| Clinical endpoint | HR (95% CI) |
| --- | --- |
| | Univariable model |
| **Any prostate cancer** | |
| MVP | 1.77 (1.72 - 1.82) |
| COSM | 1.90 (1.34 - 2.42) |
| ProtecT | 1.21 (0.92 - 1.56) |
| African | 2.50 (2.22 - 2.89) |
| Asian | 1.87 (1.14 - 2.91) |
| **Clinically significant prostate cancer** | |
| COSM | 1.74 (1.37 - 2.25) |
| ProtecT | 1.14 (0.81 - 1.58) |
| African | 2.83 (2.29 - 3.37) |
| Asian | 1.46 (0.71 - 2.60) |
| **Metastatic prostate cancer** | |
| MVP | 1.42 (1.30 - 1.56) |
| **Fatal prostate cancer** | |
| MVP | 1.56 (1.26 - 1.86) |
| COSM | 1.62 (0.92 - 2.55) |

**Supplemental Table 3: Association of family history with prostate cancer outcomes in MVP and PRACTICAL cohorts.** Data are HR (95% CI) for prostate cancer outcomes associated with reporting at least one first-degree family member with prostate cancer. Abbreviations: CI, confidence interval; COSM, Cohort of Swedish Men; HR, hazard ratio; MVP, Million Veteran Program; PRACTICAL, Prostate Cancer Association Group to Investigate Cancer Associated Alterations in the Genome; ProtecT, Prostate Testing for Cancer and Treatment.

| Predictor | Coefficient (95% CI) |
| --- | --- |
| $PHS_{601}$ | 1.09746 (1.0856 - 1.1106) |
| Family history | 0.48881 (0.4640 - 0.5100) |
| PC1 | -0.00277 (-0.0053 - -0.0005) |
| PC2 | 0.00069 (-0.0035 - 0.0045) |

**Supplemental Table 4: Coefficients in final P-CARE model.** Estimates of coefficients from LASSO-trained PHS using age at diagnosis of prostate cancer as time to event and $PHS_{601}$, family history, and the first 2 principal components of genetic ancestry as predictors. Abbreviations: CI, confidence interval; PHS, polygenic hazard score; PC, principal component.

| Clinical endpoint | N | HR (95% CI) | | | | |
|---|---|---|---|---|---|---|
| | | $HR_{SD}$ | $HR_{80/20}$ | $HR_{20/50}$ | $HR_{80/5}$ | $HR_{95/5}$ |
| **Any prostate cancer** | | | | | | |
| All | 585,418 | 2.04 (1.99 - 2.08) | 6.33 (5.95 - 6.71) | 0.43 (0.42 - 0.45) | 2.75 (2.66 - 2.84) | 4.09 (3.89 - 4.29) |
| African | 105,014 | 1.95 (1.85 - 2.05) | 5.83 (5.02 - 6.80) | 0.49 (0.45 - 0.52) | 2.82 (2.60 - 3.06) | 3.75 (3.36 - 4.19) |
| European | 420,722 | 2.05 (1.98 - 2.12) | 5.77 (5.32 - 6.22) | 0.43 (0.42 - 0.45) | 2.49 (2.38 - 2.58) | 3.81 (3.58 - 4.02) |
| American | 50,590 | 2.04 (1.86 - 2.26) | 5.56 (4.37 - 7.05) | 0.44 (0.39 - 0.49) | 2.45 (2.16 - 2.79) | 3.73 (3.12 - 4.45) |
| East Asian | 9,092 | 2.11 (1.55 - 2.85) | 5.81 (2.75 - 11.07) | 0.45 (0.31 - 0.61) | 2.42 (1.67 - 3.45) | 3.88 (2.21 - 6.56) |
| **Metastatic prostate cancer** | | | | | | |
| All | 585,418 | 2.05 (1.93 - 2.16) | 6.50 (5.50 - 7.38) | 0.43 (0.40 - 0.46) | 2.78 (2.54 - 2.99) | 4.17 (3.68 - 4.59) |
| African | 105,014 | 1.89 (1.65 - 2.19) | 5.41 (3.72 - 7.64) | 0.51 (0.44 - 0.58) | 2.69 (2.17 - 3.38) | 3.54 (2.70 - 4.75) |
| European | 420,722 | 1.94 (1.78 - 2.14) | 5.07 (4.07 - 6.43) | 0.46 (0.41 - 0.51) | 2.32 (2.07 - 2.63) | 3.45 (2.91 - 4.12) |
| American | 50,590 | 2.05 (1.41 - 2.61) | 5.83 (2.29 - 10.13) | 0.45 (0.33 - 0.68) | 2.48 (1.54 - 3.35) | 3.83 (1.88 - 5.94) |
| East Asian | 9,092 | 3.13 (0.58 - 8.53) | 61.89 (0.30 - 143.86) | 0.53 (0.09 - 1.82) | 4.27 (0.54 - 12.89) | 17.94 (0.38 - 50.76) |
| **Fatal prostate cancer** | | | | | | |
| All | 585,418 | 1.95 (1.76 - 2.15) | 5.71 (4.33 - 7.30) | 0.45 (0.41 - 0.52) | 2.59 (2.22 - 2.97) | 3.77 (3.05 - 4.57) |
| African | 105,014 | 1.62 (1.16 - 2.12) | 3.78 (1.47 - 7.06) | 0.61 (0.45 - 0.86) | 2.15 (1.25 - 3.19) | 2.67 (1.33 - 4.41) |
| European | 420,722 | 1.91 (1.63 -2.18) | 4.93 (3.28 - 6.68) | 0.47 (0.40 - 0.57) | 2.28 (1.85 - 2.68) | 3.37 (2.47 - 4.26) |
| American | 50,590 | 2.16 (0.95 - 3.91) | 7.70 (0.89 - 26.12) | 0.49 (0.21 - 1.06) | 2.67 (0.94 - 5.51) | 4.51 (0.91 - 12. 09) |
| East Asian | 9,092 | 8.91E60 (0.93 - 9.39E40) | 1.67E142 (0.85 - 1.91E95) | 1.46E113 (2.62E-37 - 1.09) | 1.73E75 (0.92 - 3.96E49) | 7.19E112 (0.88 - 7.36E75) |

**Supplemental Table 5: Association of P-CARE model with prostate cancer outcomes in genetic ancestry strata in MVP.** Association of P-CARE quantiles with any, metastatic, and fatal prostate cancer in genetic ancestry subgroups within MVP. Abbreviations: CI, confidence interval; HR, hazard ratio; MVP, Million Veteran Program; P-CARE, Prostate CAncer Risk and Evaluation.

| Clinical endpoint | N | HR (95% CI) | | | |
|---|---|---|---|---|---|
| | | $HR_{80/20}$ | $HR_{20/50}$ | $HR_{80/50}$ | $HR_{95/50}$ |
| **Any prostate cancer** | | | | | |
| All | 585,418 | 6.33 (5.95 - 6.71) | 0.43 (0.42 - 0.45) | 2.75 (2.66 - 2.84) | 4.09 (3.89 - 4.29) |
| American Indian or Alaska Native | 5507 | 5.99 (2.92 - 10.71) | 0.45 (0.34 - 0.61) | 2.56 (1.75 - 3.66) | 3.91 (2.27 - 6.17) |
| Asian | 6210 | 8.03 (2.71 - 20.08) | 0.40 (0.23 - 0.61) | 2.80 (1.66 - 4.63) | 4.88 (2.16 - 9.64) |
| Black or African American | 101,920 | 5.83 (5.05 - 6.83) | 0.48 (0.44 - 0.52) | 2.80 (2.58 - 3.04) | 3.72 (3.33 - 4.17) |
| Hispanic White | 26,037 | 5.07 (3.36 - 7.27) | 0.47 (0.39 - 0.56) | 2.32 (1.88 - 2.81) | 3.47 (2.59 - 4.57) |
| Native Hawaiian or Pacific Islander | 3042 | 6.94 (1.42 - 16.58) | 0.47 (0.26 - 0.85) | 2.69 (1.20 - 4.55) | 4.35 (1.30 - 9.01) |
| Non-Hispanic White | 418,387 | 5.78 (5.32 - 6.18) | 0.43 (0.41 - 0.45) | 2.49 (2.38 - 2.58) | 3.81 (3.58 - 4.02) |
| Other | 8077 | 7.58 (4.34 - 15.33) | 0.41 (0.29 - 0.52) | 2.97 (2.25 - 4.42) | 4.73 (3.17 - 8.03) |
| Unknown | 16,238 | 6.66 (4.20 - 9.56) | 0.43 (0.36 - 0.52) | 2.79 (2.16 - 3.47) | 4.28 (2.98 - 5.69) |
| **Metastatic prostate cancer** | | | | | |
| All | 585,418 | 6.50 (5.50 - 7.38) | 0.43 (0.40 - 0.46) | 2.78 (2.54 - 2.99) | 4.17 (3.68 - 4.59) |
| American Indian or Alaska Native | 5507 | 1.93E71 (0.15 - 390.02) | 0.55 (0.06 - 2.47) | 9.45E36 (0.36 - 23.33) | 5.31E53 (0.24 - 87.63) |
| Asian | 6210 | ∞ (0.17 - 872.47) | 0.79 (0.04 - 2.62) | 4.51E176 (0.39 - 31.10) | 1.38E255 (0.25 - 198.21) |
| Black or African American | 101,920 | 5.52 ((3.71 - 8.29) | 0.50 (0.42 - 0.58) | 2.70 (2.16 - 3.47) | 3.56 (2.66 - 4.91) |
| Hispanic White | 26,037 | 3.93 (1.12 - 8.24) | 0.57 (0.36 - 0.95) | 1.98 (1.06 - 3.01 | 2.79 (1.09 - 5.05) |
| Native Hawaiian or Pacific Islander | 3042 | 1.02E108 (0.08 - 9.70E48) | 0.76 (1.05E-19 - 3.37) | 2.43E55 (0.26 - 1.13E26) | 2.87E81 (0.13 - 9.44E37) |
| Non-Hispanic White | 418,387 | 5.16 (4.15 - 6.57) | 0.46 (0.41 - 0.50) | 2.34 (2.09 - 2.66) | 3.49 (2.96 - 4.18) |
| Other | 8077 | 18.10 (0.62 - 95.88) | 0.51 (0.12 - 1.24) | 3.78 (0.77 - 11.28) | 7.99 (0.69 - 30.46) |
| Unknown | 16,238 | 6.94 (1.89 - 16.69) | 0.47 (0.28 - 0.74) | 2.74 (1.41 - 4.53) | 4.30 (1.63 - 8.51) |
| **Fatal prostate cancer** | | | | | |
| All | 585,418 | 5.71 (4.33 - 7.30) | 0.45 (0.41 - 0.52) | 2.59 (2.22 - 2.97) | 3.77 (3.05 - 4.57) |
| American Indian or Alaska Native | 5507 | 1.95E45 (0.15 - 1.85E40) | 0.74 (4.45E-7 - 5.30) | 1.55E24 (0.29 - 3.52E21) | 1.01E37 (0.20 - 2.32E30) |
| Asian | 6210 | 3.93E305 (1.00 - 1.56E180) | 0.28 (2.72E-69 - 1.00) | 6.12E156 (1.00 - 1.54E94) | 9.67E216 (1.00 - 1.36E140) |
| Black or African American | 101,920 | 3.88 (1.35 - 7.47) | 0.61 (0.44 - 0.88) | 2.16 (1.19 - 3.30) | 2.70 (1.25 - 4.58) |
| Hispanic White | 26,037 | 9.11 (0.34 - 44.08) | 0.57 (0.17 - 1.67) | 2.64 (0.57 - 7.09) | 4.75 (0.44 - 17.74) |
| Native Hawaiian or Pacific Islander | 3042 | 1.84E92 (0.05 - 1.10E93) | 1.05 (2.32E-16 - 4.17) | 1.99E45 (0.20 - 1.18E46) | 1.38E69 (0.09 - 8.24E69) |
| Non-Hispanic White | 418,387 | 5.02 (3.39 - 6.89) | 0.48 (0.40 - 0.57) | 2.31 (1.89 - 2.73) | 3.41 (2.53 - 4.36) |
| Other | 8077 | 5.86E126 (0.01 - 1.88E50) | 1.03 (7.48E-4 - 6.95) | 2.31E67 (0.10 - 1.16E27) | 2.90E97 (0.04 - 2.30E40) |
| Unknown | 16,238 | ∞ (0.32 - 83.89) | 0.63 (0.14 - 1.67) | 3.62E199 (0.54 - 11.32) | 2.97E285 (0.42 - 28.18) |

**Supplemental Table 6:.** Association of P-CARE quantiles with any, metastatic, and fatal prostate cancer in self-reported race and ethnicity subgroups within MVP. Abbreviations: CI, confidence interval; HR, hazard ratio; MVP, Million Veteran Program; P-CARE, Prostate CAncer Risk and Evaluation.

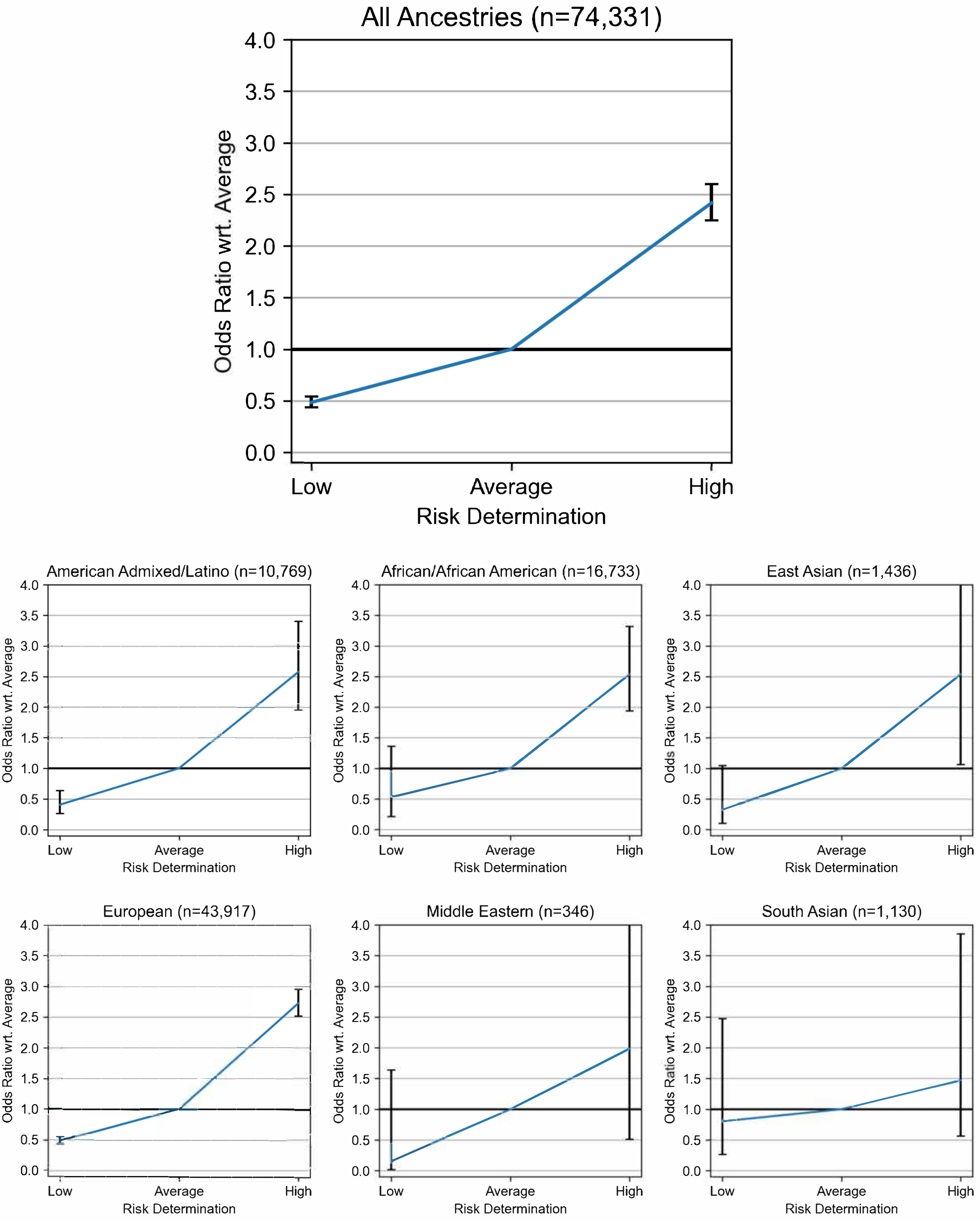
All Ancestries (n=74,331)
Odds Ratio wrt. Average
Risk Determination
Low
Average
High
American Admixed/Latino (n=10,769)
African/African American (n=16,733)
East Asian (n=1,436)
European (n=43,917)
Middle Eastern (n=346)
South Asian (n=1,130)

**Supplemental Figure 1: Odds of prostate cancer in All of Us Research Program by P-CARE category.** Shown are the odds ratios for an individual to be diagnosed with prostate cancer in the low and high P-CARE categories, relative to the average P-CARE category, derived from logistic regression models controlling for age. Error bars correspond to the 95% confidence intervals, and the shown ancestries are the predictions provided by All of Us. Abbreviations: P-CARE, Prostate CAncer Risk and Evaluation.
