## Supplemental File 2 for "From a genomic risk model to clinical trial implementation in a learning health system: the ProGRESS Study"

U.S. Department of Veterans Affairs
Veterans Health Administration
*Boston Healthcare System*

ProGRESS
PROSTATE CANCER, GENETIC RISK & EQUITABLE SCREENING STUDY

Participant Information:

First Name | Last Name | DOB | Sex | Date

**Result Summary: High risk for prostate cancer. Screening <u>strongly</u> recommended.**
***OR***
***Result Summary: Average risk for prostate cancer.*** **Shared decision making recommended.**
***OR***
***Result Summary: Low risk for prostate cancer.*** **Screening may not be recommended.**

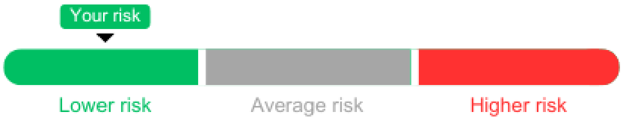


Based on analysis of your genetic sample, you have a [*insert high OR average OR low*] genetic risk of developing prostate cancer at some point in your life. These results do not indicate that you have prostate cancer now or will definitely develop prostate cancer in the future. This test does not evaluate or report on all possible genetic results related to prostate cancer. There are steps you and your healthcare team can take to prevent development of advanced prostate cancer or diagnose and treat it early.

| | |
|---|---|
| **Hereditary Prostate Cancer Panel Result** | **Positive**<br>The following pathogenic or likely pathogenic variant(s) has been identified: [INSERT *GENE NAME]*-associated with an increased risk to develop [INSERT NAME OF CONDITION]. See details below.<br>OR<br>**Negative**<br>No pathogenic or likely pathogenic variants identified. |
| **P-CARE Result** | **High Risk [OR] Average Risk [OR] Low Risk**<br>Prostate CAncer integrated Risk Evaluation (P-CARE) is a prostate cancer risk model that includes a polygenic risk score and the family history information you shared with us to estimate your risk to develop prostate cancer. You [reported OR did not report] that you have first-degree family member(s) (father, brothers, sons) who have been diagnosed with prostate cancer. |

Based on your results, your chance for developing prostate cancer by age 80 is:

37%

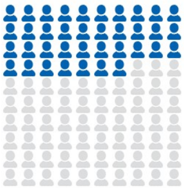

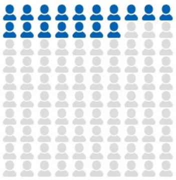

The average chance for developing prostate cancer by age 80 is:

12-16%

#### Next Steps

- [*If high risk*] You should share this result with your primary care provider to discuss ways to lower your risk of developing prostate cancer. [*If average risk*] You should share this result with your primary care provider and discuss the risks and benefits of prostate cancer screening to determine if screening is right for you. Screening is done by a blood test called a prostate-specific antigen (PSA) test. [*If low risk*] You should share this result with your primary care provider and discuss the risks and benefits of prostate cancer screening. Based on your results, the risks of prostate cancer screening may outweigh the benefits.
- [*If high risk]* Based on your results, it is <u>strongly</u> recommended you have screening for prostate cancer. The goal of screening is to detect which patients may develop aggressive prostate cancer, so they can be diagnosed and treated earlier.
- Prostate cancer screening is done by a blood test, called a prostate-specific antigen (PSA) test. If your PSA result is abnormal, a doctor may recommend additional testing, such as a prostate MRI.
- Your genetic results will be entered into your VA medical record and shared with your primary care provider.

### Prostate Cancer Information

#### What is prostate cancer?

Prostate cancer is a disease in which cells in the prostate grow out of control. Prostate cancer is one of the most common cancers in men. All men are at risk for prostate cancer, and the older a man gets, the larger the chance of getting prostate cancer. Prostate cancer can be slow-growing or aggressive. Slow-growing cancer sits in the prostate without spreading, while aggressive prostate cancer grows more quickly and can spread to other parts of the body and even cause death.

#### What is a polygenic risk score?

A polygenic risk score is a genetic test that takes into account hundreds of genetic variants to estimate the risk to develop a disease. People with a high score are more likely to develop prostate cancer, while people with a low score are less likely to develop prostate cancer.

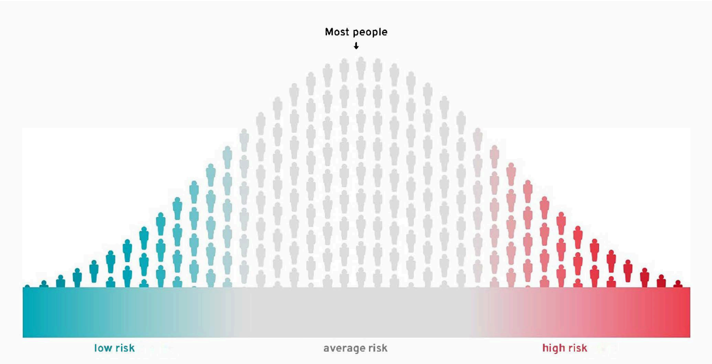


Lifetime risk of prostate cancer by P-CARE risk category*

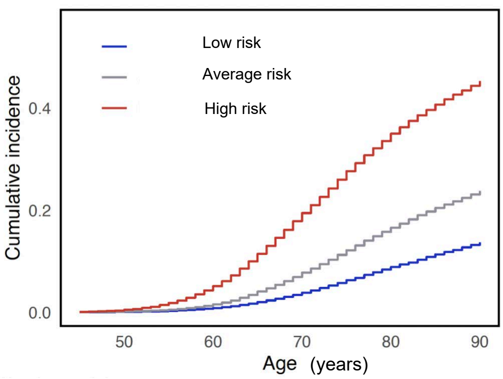


*Based on risk among participants in the Million Veteran Program

#### What is the P-CARE model?

The Prostate CAncer integrated Risk Evaluation (P-CARE) model combines your polygenic risk score with information from your family history and genetic background. It predicts your risk of prostate cancer. Having a high P-CARE score means you might be at high risk for developing prostate cancer in the future, including metastatic and lethal prostate cancer.

#### What are other risk factors for prostate cancer?

Risk factors for prostate cancer include age, race/ethnicity, smoking history, diet, and environmental and other exposures. The P-CARE model includes genetic factors but does not include environmental or healthcare factors.

#### How do race and ethnicity impact prostate cancer risk?

Black/African-American men have double the risk of developing prostate cancer compared to White men. Some of this increased risk may be due to genetic factors, access to healthcare, socioeconomic status, and lifestyle factors.

#### How is prostate cancer detected and treated?

The prostate specific antigen (PSA) test is a blood test that is used to screen for prostate cancer. If a patient's PSA result is abnormal, it is usually repeated to confirm the result. If the PSA remains abnormal, a doctor may recommend a prostate MRI to look for signs of cancer. Some patients may then require a prostate biopsy, the standard procedure used to diagnose prostate cancer.

Some prostate cancers do not need to be diagnosed or treated. If a prostate cancer is slow growing, it is safe for it to be monitored over time. More aggressive prostate cancers that are detected at an early stage may be cured with surgery and/or radiation therapy. Late-stage cancers may require treatment with medical therapy

#### What does this mean for my family members?

Your family members (parents, siblings, children) may consider speaking to their own doctors about genetic testing and/or screening for prostate cancer based on their own personal risk factors.

BROAD INSTITUTE | CLINICAL RESEARCH SEQUENCING PLATFORM

ProGRESS
PROSTATE CANCER, GENETIC RISK & EQUITABLE SCREENING STUDY

#### Patient

**Patient Name:** Jane Doe
**DOB:** 04/25/1993 Age: 30
**Sex:** Female
**MRN:** 00006 Accession **ID:**
Sample 6 Family ID:

#### Specimen

**Specimen Type:** Bodily Fluid:Saliva
**Collection Date:** 01/01/2023
**Received Date:** 01/03/2023 Report
**Date:** Not yet approved

#### Ordering Physician

**Ordering Physician:** Rosalind Franklin
**Ordering Physician Address:** 10001 1st Ave, Cambridge, MA 12345

#### Indication for Testing

Hereditary Prostate Cancer Screening

#### Test Name

Genome Sequencing - Hereditary Prostate Cancer Panel (12 genes), Broad Institute

#### Test Result

Positive Result A pathogenic/likely pathogenic variant was detected.

| GENE | VARIANT | VARIANT CLASSIFICATION | CONDITION | INHERITANCE PATTERN | ZYGOSITY | PARENTAL ORIGIN |
|---|---|---|---|---|---|---|
| BRCA2 | c.9076C>T p.Gln3026Ter | Pathogenic | Hereditary breast and ovarian cancer | Autosomal dominant | Heterozygous | Not Tested |

A single variant was identified in this test. One [pathogenic/likely pathogenic] variant was identified in [gene], which is associated with [disease], with autosomal [recessive/dominant] inheritance. Additional information about these findings is included in Variant Details.

As part of the ProGRESS study, some individuals may have received polygenic risk assessment in conjunction with this monogenic gene panel test. Please refer to the ProGRESS Study Result Summary for prostate cancer screening recommendations.

#### Recommendations

Genetic counseling is available to help individuals understand these results and address any questions. You may contact the ProGRESS genetic counselor at. For further assistance in locating nearby genetic counseling services, please contact your provider or find a genetic counselor through the National Society of Genetic Counselors (https://findageneticcounselor.nsgc.org/). For questions about this report and interpretation please contact.

Cancer screening recommendations should factor in personal and family history, genetic and non-genetic risk factors, and population-based guidelines. Screening recommendations, including high risk screening, can be found through the National Comprehensive Cancer Network at www.nccn.org. Work with your healthcare team to determine what cancer screening and/or management is best for you.

Individuals who meet genetic testing criteria for hereditary cancer risk based on personal and/or family history may require additional testing. To determine the most appropriate testing recommendations, contact your health care team or high-risk specialists. These results should be interpreted in the context of the patient's medical and family history. Please note that variant classification may change over time if more information becomes available. Testing limitations are detailed below.



DRAFT

#### Patient

**Patient Name:** Jane Doe
**DOB:** 04/25/1993
**MRN:** 00006
**Sex:** Female
**Age:** 30
**Accession ID:** Sample 6

#### Variant Details

**A pathogenic variant in BRCA2 was identified**

**Gene and Disease Summary**

The BRCA2 gene encodes a tumor suppressor protein that is involved in maintenance of genome stability, especially the homologous recombination pathway for double-strand DNA repair. Pathogenic variants in BRCA2 are associated with autosomal dominant hereditary breast and ovarian cancer (HBOC) syndrome. Pathogenic variants are associated with greatly elevated risk of breast cancer (especially ER + cancer) and ovarian cancer and a small but significant elevated risk of pancreatic and prostate cancers. There is also limited evidence for association with melanoma and leukemia.

**Variant Summary**

BRCA2 NM_000059.3: c.9076C>T (NP_000050.2: p.Gln3026Ter) *[chr13:32954009 (GRCh37)]*

This variant introduces a premature termination signal at codon 3026 in the BRCA2 protein, which is predicted to lead to a truncated or absent protein. Heterozygous loss-of-function of the BRCA2 gene is an established disease mechanism in hereditary breast and ovarian cancer (PMID: 20104584). This variant is absent from large population studies. This variant has been reported in individuals with clinical features of hereditary breast and ovarian cancer (PMID: 19016756, 12161607, 25863477, 26187060, 28008555). ClinVar contains an entry for this variant (Variation ID: 38207). For these reasons this variant has been classified as Pathogenic (ACMG/AMP Criteria applied: PVS1_strong, PM2, PM1_moderate, PS4_moderate).

DRAFT

#### Patient

**Patient Name:** Jane Doe | **DOB:** 04/25/1993 | **MRN:** 00006
**Sex:** Female | **Age:** 30 | **Accession ID:** Sample 6

#### Test Methodology

Genomic DNA extracted from blood or saliva is fragmented, adapter ligated, and barcoded. Library fragments are sequenced (2x150 base paired end) using Sequencing-By-Synthesis (SBS) chemistry and the Illumina NovaSeq sequencer. Sequence data are aligned to the human reference sequence (GRCh37 assembly; hg19) after discarding low quality sequences. Illumina's DRAGEN (Dynamic Read Analysis for GENomics) platform is used for demultiplexing, read mapping, genome alignment, read sorting, duplicate marking, and variant calling. The DRAGEN pipeline generates quality metrics. Specimen data must meet the following thresholds: ≥ 90% of bases at or greater than 20X and mean coverage ≥ 30X. Other quality metrics that are reviewed are percent contamination (≤ 2.5%), percent mapped (≥ 75%), and percent callability (≥ 95%). The DRAGEN pipeline also generates a cram file (compressed BAM file) and multiple vcf (Variant Call Format) files. A concatenated vcf file containing small variants (single nucleotide variants and small insertions/deletions) and copy number variants is used as input to the Fabric Genomics platform, which utilizes the ACE pipeline for small variant identification, filtering, and classification. ACE provides preliminary auto-classification of small variants based on the codable ACMG/AMP sequence variant interpretation evidence codes (Richards et al 2015. PMID: 25741868). Copy number variants that do not pass DRAGEN quality metrics and are smaller than 1kb are removed, and the remaining copy number variants are reviewed manually. Copy number variants <1kb may be unmasked to identify a second variant if there is a single pathogenic or likely pathogenic variant identified in a gene associated with autosomal recessive disease. Initial manual review of auto-applied and manually curated variant evidence is performed by the Fabric Genomics' Clinical Services team with secondary review and sign out by the Clinical Research Sequencing Platform's clinical team. All variants located in coding and flanking intronic regions are examined as well as any variant classified in ClinVar as P/LP within introns, UTRs, and 5kb upstream and downstream of the gene transcript listed in the appendix of this report. Variants are reported using HGVS nomenclature (www.hgvs.org/mutnomen) and classified according to ACMG/AMP guidelines.

Only variants classified as Pathogenic or Likely Pathogenic in genes recommended for return by ACMG are reported (see Appendix for gene list). Variants in autosomal recessive genes are only reported if Pathogenic or Likely Pathogenic variants are found in both copies of the gene. If it is not possible to determine if two variants are on the same copy or different copies of a gene, both variants will be reported. In addition, the HFE p.C282Y variant in the homozygous state is the only variant reported in the HFE gene. For genes with multiple associated phenotypes, only variants classified as Pathogenic or Likely Pathogenic for the phenotypes deemed reportable as secondary findings are returned (Miller et al 2022. PMID: 35802134). Variants of uncertain significance are not reportable in any gene and will not be returned. Copy number variants are reported in the context of their impact to panel genes only (see Test Limitations section below).

Larger reportable copy number events are confirmed using an orthogonal microarray-based method. This confirmation test is sent out to GeneDx, LLC. (CLIA: 21D0969951) where they perform the ExonArrayDx. "This array is a GeneDx-designed oligonucleotide microarray constructed for identification of exonic deletions or duplications within or encompassing a targeted gene. Location of ExonArrayDx probes is based on human genome build hg19 (GRCh37). Analysis of array data is performed using Genomic Workbench software (Agilent Technologies). The array design is optimized based on its performance and on newly published gene information." (GeneDx, LLC. ExonArrayDx test information sheet)

#### Test Limitations

Please note that certain variants including short tandem repeats, structural rearrangements (e.g. translocations, inversions, gene conversion events, etc), and large chromosomal events (e.g. aneuploidy) as well as variants in highly repetitive regions (e.g. centromeric and telomeric), in the mitochondrial genome, and regions with low coverage, mapping quality, and base quality are currently not reliably detected by short-read whole genome sequencing and not evaluated as part of this test. Variants of uncertain significance are not reported nor are variants that do not meet the guidelines as reportable secondary findings noted above. All interpretations are based on currently available data. However, these interpretations may change as new information becomes available (see Methodology). A new report may be released to the ordering provider if new information generates a reportable update. There may also be limited detection of low-level mosaicism. The sequencing data is limited to the tissue source used for DNA extraction. Variants in genes outside of the genes included on this panel were not analyzed. In addition, the small variants (single



DRAFT

#### Patient

**Patient Name:** Jane Doe | **DOB:** 04/25/1993 | **MRN:** 00006

**Sex:** Female | **Age:** 30 | **Accession ID:** Sample 6

#### Test Limitations (CONT.)

nucleotide variant and small insertions or deletions) are not confirmed using a second testing method, though a secondary fingerprinting assay is used to confirm sample swaps have not occurred during sample processing. Because this panel analysis is performed from whole genome sequencing data, it is possible that a copy number event that extends outside a panel gene is detected. This report only includes information about the copy number event's impact on the genes designated on this panel. This analysis does not assess the full extent of a variant outside of a panel gene. Furthermore, resolution of exact breakpoints of copy number events from short read sequencing data may not be possible and additional clinical confirmatory studies are indicated in those cases. This information could impact the predicted pathogenicity of a copy number variant depending on the exact disruption point within the gene.

#### Regulatory Disclosures

This test was performed by the Clinical Research Sequencing Platform (CRSP), LLC. (320 Charles Street, Cambridge, MA 02141; CLIA: 22D2055652). CRSP, LLC. is authorized under the Clinical Laboratory Improvement Amendments (CLIA) to develop and perform high complexity clinical laboratory testing. This test was developed and its performance characteristics determined by CRSP, LLC. The U.S. Food and Drug Administration (FDA) has not approved or cleared this test; however, FDA clearance or approval is not currently required for clinical use.

Clinical Laboratory Director: Heidi Rehm, PhD, FACMG

CLIA: 22D2055652, CAP: 8707596

Commonwealth of Massachusetts Clinical Laboratory License: 5347

DRAFT

#### Patient

**Patient Name:** Jane Doe **DOB:** 04/25/1993 **MRN:** 00006

**Sex:** Female **Age:** 30 **Accession ID:** Sample 6

### APPENDIX

#### Genes Evaluated

*ATM, BRCA1, BRCA2, CHEK2, EPCAM, HOXB13, MLH1, MSH2, MSH6, PALB2, PMS2, TP53*

DRAFT

BROAD INSTITUTE | CLINICAL RESEARCH SEQUENCING PLATFORM

Sample ID: 10MX1002

### ProGRESS P-CARE Report

| PATIENT INFORMATION | | REFERRING PROVIDER | | SPECIMEN | |
|---|---|---|---|---|---|
| Patient Name: | Johan Smith | Provider Name: | Dr. Jason Vassy | Report Type: | Final |
| Date of Birth: | 01/01/1990 | Referring Facility: | VA Boston Healthcare System | Collected: | 10/01/2020 |
| Patient ID: | e4567 | Test Performed: | Polygenic Risk Evaluation | Received: | 10/01/2020 |
| Sample ID: | 10MX1002 | Indication: | N/A | Report Date: | 11/01/2020 |
| Accession ID: | SM-XXXXX | | | Report Updated Date: | 11/01/2021 |
| Site Sample ID: | 12345678 | | | Material Type: | DNA |
| | | | | Material Source: | Blood |

#### Results Summary

In this patient the Prostate CAncer integrated Risk Evaluation (P-CARE) result for prostate cancer was determined to be:

**HIGH RISK FOR PROSTATE CANCER** based on combined genetic score and family history
[*OR*]
**AVERAGE RISK FOR PROSTATE CANCER** based on combined genetic score and family history
[*OR*]
**LOW RISK FOR PROSTATE CANCER** based on combined genetic score and family history

*See detailed results for a description for how this risk was determined.

#### Detailed Results

**Prostate Cancer Risk: HIGH RISK [*OR*] AVERAGE RISK [*OR*] LOW RISK**

1. Prostate cancer is a disease of abnormal cell growth in the prostate. It is the most common non-skin cancer in men and the 2nd leading cause of cancer death in men. All men are at risk for prostate cancer; however, the risks for developing and dying from prostate cancer increase with age, with the highest incidence observed in men over 65. Prevalence of prostate cancer varies among different ethnic and racial groups, with the highest prevalence observed in African-American men. Prostate cancer is often asymptomatic in its early stages but can be associated with bone pain if it spreads to other parts of the body.

2. A [*high OR average OR low*] risk P-CARE result for prostate cancer was found in this individual. P-CARE is a prostate cancer risk model that combines a polygenic score, genetic ancestry, and family history of prostate cancer. Higher P-CARE results are associated with higher risk of developing any prostate cancer, metastatic prostate cancer, and lethal prostate cancer. For this report, P-CARE results are categorized as low, average, or high based on their association with metastatic prostate cancer. A high P-CARE result is associated with at least a 1.5-fold hazard (50% increased hazard) for developing metastatic prostate cancer relative to median P-CARE values (40th-60th percentile). A low P-CARE result is associated with a 0.75-fold hazard (25% lower hazard) for developing metastatic prostate cancer relative to median P-CARE values (40th-60th percentile). All other P-CARE results are categorized as average risk.

3. Factors including monogenic disease risk, family history, and other clinical measures can have an impact on the individual's overall (absolute) risk and should be considered. This result should be interpreted in the context of the patient's medical history, family history, and racial/ethnic background.

### Methodology

#### Blended genome-exome

Data is generated from blending whole exome sequencing data at a mean target depth of >85X with whole genome sequencing data at a mean target depth of 2.5X coverage. The input into the cBGE process is genomic DNA that was extracted from either blood, saliva, or buccal swabs. gDNA was enzymatically fragmented, adapter ligated, and barcoded to create a single PCR free whole genome. An aliquot from the genome was taken through PCR amplification and exome selection. A blend of the PCR free genome and whole exome was created and sequenced to deliver an optimal coverage balance. Library fragments were sequenced (2x150 base paired end) using Sequencing-By-Synthesis (SBS) chemistry on the Illumina NovaSeq X Plus sequencer. Sequencing data were aligned to the GRCh38 assembly after discarding low quality sequences. Illumina's DRAGEN (Dynamic Read Analysis for GENomics) platform was used for demultiplexing, read mapping, genome alignment, read sorting, duplicate marking, and variant calling. The DRAGEN pipeline generates the above quality metrics, a CRAM file, and a VCF (Variant Call Format) file that contains SNPs/indels with a target region of +/-16 base pairs into the introns. Detailed methods are available upon request. Assay limitations for Germline Analysis are below. Highly accurate variant calling is performed over the exome using the Illumina DRAGEN pipeline (ADD reference). The combined high coverage exome and lower coverage whole genome is also imputed with GLIMPSE2 using a reference panel based on data from the 1000 Genomes Project. Polygenic Risk Scores are calculated using the high quality exome variant calling results where available, and the GLIMPSE2 imputed genotypes for other sites.

#### P-CARE model

The Prostate CAncer integrated Risk Evaluation (P-CARE) model is an updated version of an earlier model described in Pagadala JNCI 2023 and was developed among 646,991 participants of the Million Veteran Program (18.9% African, 70.6% European, 8.8% American, 1.6% East Asian) and validated among 18,457 participants in the PRACTICAL Consortium. Validation information may be insufficient or not available for populations of other descent. Variants considered for inclusion in the PHS included variants from a prior PHS, variants identified as prostate cancer susceptibility loci in multi-ancestry genome-wide association studies, variants identified as susceptibility loci for benign elevation of prostate-specific antigen (PSA) or benign prostatic hypertrophy (BPH), variants identified as prostate cancer susceptibility loci in men of African ancestry in a genome-wide meta-analysis, and variants identified as susceptibility loci for prostate cancer in a genome-wide multi-ancestry meta-analysis. The final P-CARE model consists of a polygenic hazard score of 601 variants (coefficient 1.097), patient-reported history of prostate cancer in at least one first-degree family member (coefficient 0.4889), and the first 2 principal components of genetic ancestry derived from 2,309 ancestry-informative markers (coefficients -0.0028 and 0.001). In the Million Veteran Program, a P-CARE result above the 80th percentile was associated with a hazard ratio of 2.75 (2.66-2.84), 2.78 (2.54 - 2.99), and 2.59 (2.22 - 2.97) for any prostate cancer, clinically significant prostate cancer, and metastatic prostate cancer, respectively, compared to the median. A P-CARE result below the 20th percentile was associated with a hazard ratio of 0.43 (0.42 - 0.45), 0.43 (0.40 - 0.46), and 0.45 (0.41 - 0.52) for any prostate cancer, clinically significant prostate cancer, and metastatic prostate cancer, respectively, compared to the median.

#### Limitations

- A P-CARE result is neither deterministic nor diagnostic. Some people with a 'high risk' result will never develop prostate cancer, while others with a 'low risk' result still have a risk of developing prostate cancer. Therefore, this test is not intended to diagnose a disease or to make surgical or pharmacological intervention decisions. This test does not tell a patient anything about their current state of health and should not substitute for regular visits to the doctor. Any diagnostic or treatment decisions should be based on additional testing and/or other information that is managed by a healthcare provider.
- This test detects genetic variants that are predefined. This test does not evaluate or report on all possible genetic variation related to a given disease. The test used for the P-CARE model will not detect novel or rare genetic variants and will not rule out the presence of these additional genetic variants related to a disease.
- The predefined list of genetic variants tested in this assay may be different from another institution or company, therefore genetic risk calculations and polygenic scores may differ if compared across different institutions or companies.
- The hazard ratio (HR) listed for each condition does not take into account other factors that may play a role in a patient's overall risk of developing a disease (e.g monogenic risk of developing the disease or environmental and lifestyle risk factors).
- Receiving genetic testing results may bring up a variety of emotions or questions for patient. Patients should speak to their doctor or healthcare provider regarding these test results and potential implications for their health and lifestyle decisions. If you have questions regarding your test results, you can contact the ProGRESS clinical study team using the instructions provided to you during the consent process.
- Although the polygenic score and P-CARE model have been developed to maximize the ability to predict risk in all ancestries, the availability of population reference data means that the score is currently most accurate for those with European genetic ancestry. Due to this population limitation, as well as assay performance and processing issues, some patients may receive a result of "Not Resulted."

Test run under the supervision of Heidi Rehm, PhD, FACMG
