## Supplementary material for "From a genomic risk model to clinical trial implementation in a learning health system: the ProGRESS Study": Acknowledgements and Funding

**ACKNOWLEDGEMENT AND FUNDING STATEMENT**

**Acknowledgements**

This publication does not represent the views of the Department of Veterans Affairs or the United States Government.

**VA Million Veteran Program Core Acknowledgement for Publications. MVP Program Office:** Sumitra Muralidhar, Ph.D., Program Director, US Department of Veterans Affairs, 810 Vermont Avenue NW, Washington, DC 20420; Jennifer Moser, Ph.D., Associate Director, Scientific Programs, US Department of Veterans Affairs, 810 Vermont Avenue NW, Washington, DC 20420; Jennifer E. Deen, B.S., Associate Director, Cohort & Public Relations, US Department of Veterans Affairs, 810 Vermont Avenue NW, Washington, DC 20420. **MVP Executive Committee:** Co-Chair: Philip S. Tsao, Ph.D., VA Palo Alto Health Care System, 3801 Miranda Avenue, Palo Alto, CA 94304; Co-Chair: Sumitra Muralidhar, Ph.D.
US Department of Veterans Affairs, 810 Vermont Avenue NW, Washington, DC 20420; J. Michael Gaziano, M.D., M.P.H., A Boston Healthcare System, 150 S. Huntington Avenue, Boston, MA 02130; Elizabeth Hauser, Ph.D., Durham VA Medical Center, 508 Fulton Street, Durham, NC 27705;, Amy Kilbourne, Ph.D., M.P.H., VA HSR&D, 2215 Fuller Road, Ann Arbor, MI 48105; Michael Matheny, M.D., M.S., M.P.H., VA Tennessee Valley Healthcare System, 1310 24^th^ Ave. South, Nashville, TN 37212;, Dave Oslin, M.D., Philadelphia VA Medical Center, 3900 Woodland Avenue, Philadelphia, PA 19104. **MVP Co-Principal Investigators:** J. Michael Gaziano, M.D., M.P.H., VA Boston Healthcare System, 150 S. Huntington Avenue, Boston, MA 02130; Philip S. Tsao, Ph.D.. VA Palo Alto Health Care System, 3801 Miranda Avenue, Palo Alto, CA 94304. **MVP Core Operations:** Jessica V. Brewer, M.P.H., Director, MVP Recruitment & Enrollment VA Boston Healthcare System, 150 S. Huntington Avenue, Boston, MA 02130; Mary T. Brophy M.D., M.P.H., Director, VA Central Biorepository, VA Boston Healthcare System, 150 S. Huntington Avenue, Boston, MA 02130; Kelly Cho, M.P.H, Ph.D., Director, MVP Phenomics Data Core, VA Boston Healthcare System, 150 S. Huntington Avenue, Boston, MA 02130; Lori Churby, B.S., Director, MVP Regulatory Affairs VA Palo Alto Health Care System, 3801 Miranda Avenue, Palo Alto, CA 94304; Scott L. DuVall, Ph.D., Director, VA Informatics and Computing Infrastructure (VINCI) VA Salt Lake City Health Care System, 500 Foothill Drive, Salt Lake City, UT 84148; Saiju Pyarajan Ph.D., Director, Data and Computational Sciences
VA Boston Healthcare System, 150 S. Huntington Avenue, Boston, MA 02130; Luis E. Selva, Ph.D., Executive Director, MVP Biorepositories, VA Boston Healthcare System, 150 S. Huntington Avenue, Boston, MA 02130; Shahpoor (Alex) Shayan, M.S., Director, MVP Recruitment and Enrollment Informatics, VA Boston Healthcare System, 150 S. Huntington Avenue, Boston, MA 02130; Stacey B. Whitbourne, Ph.D., Director, MVP Cohort Management
VA Boston Healthcare System, 150 S. Huntington Avenue, Boston, MA 02130. **MVP Coordinating Centers:** MVP Coordinating Center, Boston – J. Michael Gaziano, M.D., M.P.H.
VA Boston Healthcare System, 150 S. Huntington Avenue, Boston, MA 02130; MVP Coordinating Center, Palo Alto – Philip S. Tsao, Ph.D., VA Palo Alto Health Care System, 3801 Miranda Avenue, Palo Alto, CA 94304; MVP Information Center, Canandaigua – Brady Stephens, M.S., Canandaigua VA Medical Center, 400 Fort Hill Avenue, Canandaigua, NY 14424; Cooperative Studies Program Clinical Research Pharmacy Coordinating Center, Albuquerque – Todd Connor, Pharm.D.; Dean P. Argyres, B.S., M.S.; New Mexico VA Health Care System, 1501 San Pedro Drive SE, Albuquerque, NM 87108. **MVP Publications and Presentations Committee:** Co-Chair: Themistocles L. Assimes, M.D., Ph. D, VA Palo Alto Health Care System, 3801 Miranda Avenue, Palo Alto, CA 94304; Co-Chair: Adriana Hung, M.D.; M.P.H, VA Tennessee Valley Healthcare System, 1310 24^th^ Ave. South, Nashville, TN 37212; Co-Chair: Henry Kranzler, M.D., Philadelphia VA Medical Center, 3900 Woodland Avenue, Philadelphia, PA 19104.

**Funding**

This work was funded by the Million Veteran Program MVP022 award #I01CX001727 (PI: RLH) and MVP084 award #I01CX002635 (PI: JLV).

It was supported using resources and facilities of the Department of Veterans Affairs (VA) Informatics and Computing Infrastructure (VINCI) ORD 24-VINCI-01, including writing support from Kathryn Pridgen, under the research priority to Put VA Data to Work for Veterans (VA ORD 24-D4V). Funding for salaries includes: Department of Veterans Affairs (VISN22 Veterans Center of Excellence for Stress and Mental Health to RLH), VA Office of Research and Development (1I01CX002709, 1I01CX002622 to KNM), National Institutes of Health (R01AG050595 to RLH, K08CA215312 to KNM), the Department of Defense (DOD/CDMRP PC220521 to TMS), the Prostate Cancer Foundation (23CHAL12 to TMS, 20YOUN02 to KNM, 22CHAL02 to IPG, BSR, KNM), the Burroughs Wellcome Foundation (#1017184 to KNM), Basser Center for BRCA (KNM).

The CAP trial was funded by grants C11043/A4286, C18281/A8145, C18281/A11326, C18281/A15064; and C18281/A24432 from Cancer Research UK. The UK Department of Health, National Institute of Health Research provided partial funding. The ProtecT trial was funded by project grants 96/20/06 and 96/20/99 from the UK National Institute for Health Research, Health Technology Assessment Programme. RMM is a National Institute for Health Research Senior Investigator (NIHR202411). RMM is supported by a Cancer Research UK 25 (C18281/A29019) programme grant (the Integrative Cancer Epidemiology Programme). RMM is also supported by the NIHR Bristol Biomedical Research Centre which is funded by the NIHR (BRC-1215-20011) and is a partnership between University Hospitals Bristol and Weston NHS Foundation Trust and the University of Bristol. Department of Health and Social Care disclaimer: The views expressed are those of the author(s) and not necessarily those of the NHS, the NIHR or the Department of Health and Social Care. AV is supported by Spanish Instituto de Salud Carlos III (ISCIII) funding, an initiative of the Spanish Ministry of Economy and Innovation partially supported by European Regional Development FEDER Funds (PI22/00589, INT24/00023, DTS24/00083); and by the AECC (PRYES211091VEGA). ASK is supported by National Institutes of Health, Grant/Award Numbers: U01 - U01CA268810.

CRUK and PRACTICAL Consortium

This work was supported by the Canadian Institutes of Health Research, European Commission's Seventh Framework Programme grant agreement n° 223175 (HEALTH-F2-2009-223175), Cancer Research UK Grants C5047/A7357, C1287/A10118, C1287/A16563, C5047/A3354, C5047/A10692, C16913/A6135, and The National Institute of Health (NIH) Cancer Post-Cancer GWAS initiative grant: No. 1 U19 CA 148537-01 (the GAME-ON initiative).

We would also like to thank the following for funding support: The Institute of Cancer Research and The Everyman Campaign, The Prostate Cancer Research Foundation, Prostate Research Campaign UK (now PCUK), The Orchid Cancer Appeal, Rosetrees Trust, The National Cancer Research Network UK, The National Cancer Research Institute (NCRI) UK. We are grateful for support of NIHR funding to the NIHR Biomedical Research Centre at The Institute of Cancer Research, The Royal Marsden NHS Foundation Trust, and Manchester NIHR Biomedical Research Centre. The Prostate Cancer Program of Cancer Council Victoria also acknowledge grant support from The National Health and Medical Research Council, Australia (126402, 209057, 251533, , 396414, 450104, 504700, 504702, 504715, 623204, 940394, 614296,), VicHealth, Cancer Council Victoria, The Prostate Cancer Foundation of Australia, The Whitten Foundation, PricewaterhouseCoopers, and Tattersall’s. EAO, DMK, and EMK acknowledge the Intramural Program of the National Human Genome Research Institute for their support.

Genotyping of the OncoArray was funded by the US National Institutes of Health (NIH) [U19 CA 148537 for ELucidating Loci Involved in Prostate cancer SuscEptibility (ELLIPSE) project and X01HG007492 to the Center for Inherited Disease Research (CIDR) under contract number HHSN268201200008I]. Additional analytic support was provided by NIH NCI U01 CA188392 (PI: Schumacher).

Research reported in this publication also received support from the National Cancer Institute of the National Institutes of Health under Award Numbers U10 CA37429 (CD Blanke), and UM1 CA182883 (CM Tangen/IM Thompson). The content is solely the responsibility of the authors and does not necessarily represent the official views of the National Institutes of Health.

Funding for the iCOGS infrastructure came from: the European Community's Seventh Framework Programme under grant agreement n° 223175 (HEALTH-F2-2009-223175) (COGS), Cancer Research UK (C1287/A10118, C1287/A 10710, C12292/A11174, C1281/A12014, C5047/A8384, C5047/A15007, C5047/A10692, C8197/A16565), the National Institutes of Health (CA128978) and Post-Cancer GWAS initiative (1U19 CA148537, 1U19 CA148065 and 1U19 CA148112 - the GAME-ON initiative), the Department of Defence (W81XWH-10-1-0341), the Canadian Institutes of Health Research (CIHR) for the CIHR Team in Familial Risks of Breast Cancer, Komen Foundation for the Cure, the Breast Cancer Research Foundation, and the Ovarian Cancer Research Fund.

BPC3

The BPC3 was supported by the U.S. National Institutes of Health, National Cancer Institute (cooperative agreements U01-CA98233 to D.J.H., U01-CA98710 to S.M.G., U01-CA98216 to E.R., and U01-CA98758 to B.E.H., and Intramural Research Program of NIH/National Cancer Institute, Division of Cancer Epidemiology and Genetics).

CAPS

CAPS GWAS study was supported by the Cancer Risk Prediction Center (CRisP; www.crispcenter.org), a Linneus Centre (Contract ID 70867902) financed by the Swedish Research Council, (grant no K2010-70X-20430-04-3), the Swedish Cancer Foundation (grant no 09-0677), the Hedlund Foundation, the Soederberg Foundation, the Enqvist Foundation, ALF funds from the Stockholm County Council. Stiftelsen Johanna Hagstrand och Sigfrid Linner's Minne, Karlsson's Fund for urological and surgical research.

PEGASUS

PEGASUS was supported by the Intramural Research Program, Division of Cancer Epidemiology and Genetics, National Cancer Institute, National Institutes of Health.
